# Analysis of functional connectivity using machine learning and deep learning in multimodal data from patients with schizophrenia

**DOI:** 10.1101/2022.11.06.22282001

**Authors:** Caroline L. Alves, Thaise G. L. de O. Toutain, Joel Augusto Moura Porto, Patricia de Carvalho Aguiar, Aruane M. Pineda, Francisco A. Rodrigues, Eduardo Pondé de Sena, Christiane Thielemann

## Abstract

*Schizophrenia* is a severe mental disorder associated with persistent or recurrent psychosis, hallucinations, delusions, and thought disorders that affect approximately 26 million people worldwide, according to the World Health Organization (WHO). Several studies encompass machine learning and deep learning algorithms to automate the diagnosis of this mental disorder. Others study schizophrenia brain networks to get new insights into the dynamics of information processing in patients suffering from the condition. In this paper, we offer a rigorous approach with machine learning and deep learning techniques for evaluating connectivity matrices and measures of complex networks to establish an automated diagnosis and comprehend the topology and dynamics of brain networks in schizophrenia patients. For this purpose, we employed an fMRI and EEG dataset in a multimodal fashion. In addition, we combined EEG measures, i.e., Hjorth mobility and complexity, to complex network measurements to be analyzed in our model for the first time in the literature. When comparing the schizophrenia group to the control group, we found a high positive correlation between the left superior parietal lobe and the left motor cortex and a positive correlation between the left dorsal posterior cingulate cortex and the left primary motor. In terms of complex network measures, the diameter, which corresponds to the longest shortest path length in a network, may be regarded as a biomarker because it is the most important measure in a multimodal fashion. Furthermore, the schizophrenia brain networks exhibit less segregation and lower distribution of information. As a final result, EEG measures outperformed complex networks in capturing the brain alterations associated with schizophrenia. As a result, our model achieved an AUC of 100%, an accuracy of 98% for the fMRI, an AUC of 95 %, and an accuracy of 95% for the EEG data set. These are excellent classification results. Furthermore, we investigated the impact of specific brain connections and network measures for these results, which helped us better describe changes in the diseased brain.

## I. INTRODUCTION

Schizophrenia is a mental disorder that has plagued individuals for millennia and affects around 26 million people worldwide, according to the World Health Organization (WHO) [1]. Archaeologists discovered ancient Egyptian inscriptions outlining common signs of this mental disorder [2]. However, it was not until the nineteenth century that it was classified as dementia praecox by psychiatrist Emil Kraepelin, who claimed that people with this condition suffered from constant and permanent mental degeneration beginning in childhood. In 1908, the Swiss physician Eugen Bleuler dubbed this psychiatric condition *schizophrenia* [3] (SCZ), which means split mind in Greek because one of its symptoms was a loss of mind and awareness unity [4]. Other symptoms of SCZ encompass: persistent or recurring psychosis, hallucinations (mainly auditory voices), delusions, and disordered thinking [5, 6].

Despite centuries of research, it is still unknown what biologically causes schizophrenia [7]. The authors in [8] propose a functional and structural disconnection of brain networks, resulting in a dysfunctional integration of them, reflecting on numerous cognitive and behavioral symptoms of schizophrenia [9]. This large-scale disconnection is reflected in the structural and functional topology of patients with the disorder; thus, network measures have been applied to them [4]. In [10], and [11], altered small world properties on these networks are suggested using functional magnetic resonance imaging (fMRI) data, and in [12], through electroencephalogram (EEG) data, a decrease in these properties is reported. In [13], again, using electroencephalogram data and network measurements, such as cluster coefficient and the mean of the shortest paths, it was found in the networks of patients with schizophrenia a decrease in clusters and shorter paths concerning networks of healthy patients. Although the SCZ networks still present a small world topology, there is subtle randomization resulting in a disturbance in the balance of brain integration and segregation [4].

In the investigation [14], SCZ patients, compared with control patients, had lower segregation and functional connectivity in brain areas such as the bilateral fusiform gyrus, bilateral medial temporal gyrus, left supramarginal gyrus, right amygdala, and left temporal regions. In addition, [15] brought attention to the concurrent increases and decreases in Posterior Cingulate Cortex (PCC) connectivity seen in SCZ patients, but mostly reduced connectivity between PCC and brain regions linked to the Default Mode Network (DMN). The authors also discovered significant discriminative capacity using machine learning (ML) for categorizing persons with the first episode of schizophrenia compared to control participants, with an average accuracy of 72.28% in test sample data. Furthermore, [16] discovered hypoconnectivity in the DMNs in patients with SCZ in the thalamus region. Another investigation with SCZ patients [17] showed changes in the left angular gyrus brain region. The gray matter volume of this region was 14.8% smaller than the control group in the same research, indicating that this region may represent a neuroanatomical basis for the “expression of schizophrenia”. The reversal asymmetry in the inferior parietal lobe, angular gyrus, discovered by [18], indicated a significance for this area in cognitive deficiencies, language issues, and thinking disorders in SCZ. The authors of [19] also emphasize the impairment of motor cognition in SCZ patients.

These aforementioned studies show that the structure of the brains of individuals with SCZ differs from those of normal controls. Hence, it is possible to diagnose SCZ based on data collected by the EEG or experiments. Furthermore, EEG is a low-cost, widely available technology with high temporal resolution. Therefore, EEG data have been used to study brain organization [20, 21]. fMRI, in contrast, has a poor temporal resolution but a high spatial resolution, which makes it ideal for studying spatial brain dynamics [22, 23]. fMRI scans produce a set of three-dimensional images recorded over time and measure a signal (called Blood Oxygenation Level Dependent – BOLD - signal [24]). The temporal evolution of the BOLD series is called the hemodynamic response function and is determined by the pixel intensity in fMRI images [25, 26]. Each cube of an fMRI image, called voxel, which anatomically maps a position in the brain, has a BOLD time series [27].

Previous studies have evaluated the effectiveness of ML in diagnosing SCZ with supervised machine learning algorithms that distinguish between two classes, namely SCZ and control group (see Table I). In contrast to traditional statistical methods, the ML approach does not rely on prior assumptions (such as adequate distribution, independence of observations, absence of multicollinearity, and interaction problems). It is well suited to automatically analyze and capture complex nonlinear relationships in data. In addition, new methodologies, such as SHapley Additive Explanations (SHAP) values, have evolved to aid in interpreting machine learning outcomes. Any machine learning algorithm may use this statistic to detect and prioritize features [28–30].

**TABLE I:** The table comprises machine learning research that used the same database as the current study (COBRE data described in more detail in subsection II A).

| Authors | Data | Data Type | Correlation metrics | ML methods | ROIs | AUC | Acc. | Recall | Precision |
| --- | --- | --- | --- | --- | --- | --- | --- | --- | --- |
| [31] | COBRE data | fMRI | specific method | SVM | 102 | - | 0.99 | - | - |
| [32] | COBRE data | fMRI | Pearson correlation | Deep Learning combined with SVM | 116 | - | 0.92 | - | - |
| [33] | COBRE data | fMRI | correlation-based distance metric | SVM | 90 | - | 0.65 | - | - |
| [34] | COBRE data | fMRI | Pearson correlation | SVM | 27 | - | 0.80 | - | - |
| [35] | COBRE data | fMRI | Pearson correlation | SVM | 160 | - | 0.69 | 0.92 | - |

This study aims to determine whether it is feasible to automatically detect brain changes caused by SCZ while providing a biological explanation. For that, we consider the BOLD series to develop the classification method for SCZ patients, and we also test in EEG data. After the preprocessing of these two data (A), we consider as an input for the machine learning the following data abstraction levels: (B) the correlation between the EEG electrodes and fMRI regions of interest and (C) complex network measures extracted from (B). In contrast to articles in the literature that use only one of these levels of abstraction, this study uses all two levels in both data sets in a multimodal fashion for the first time in literature. In addition, we define which of these abstraction levels is the most appropriate for capturing SCZ brain changes. The SHAP value technique has also been found to be more successful than the previous research [21, 36, 37] in finding the best brain areas, connections between brain regions, and measurements of complex networks that may be utilized to evaluate the effects of the SCZ on the brain. As a final result of this research, for the first time in literature, we combine EEG measures extracted from time series and complex network measures as input for the ML method to evaluate which measure is more critical to distinguish SCZ from control patients.

## II. MATERIALS AND METHODS

In the current study, two schizophrenia datasets were used to test our general workflow: one fMRI, described in the subsection II A, and another EEG, described in the subsection II B, with different pre-processing for each data. First, the best pairwise metrics for capturing schizophrenia-induced changes in the brain are defined based on the fMRI data workflow. This metric is then validated using EEG data.

Figure 1 depicts the fMRI complete methodology workflow used and organized into three parts according to their aim, i.e., preprocessing and using the best selecting pairwise metrics (described in Figure 1-(A) and in subsection II A 1), the best brain connection (described in Figure 1-(B) and in subsection II A 2), and the best complex network measures for differentiating schizophrenia from control group (described in Figure 1-(C) and in subsection II A 3.)

**FIG. 1:**
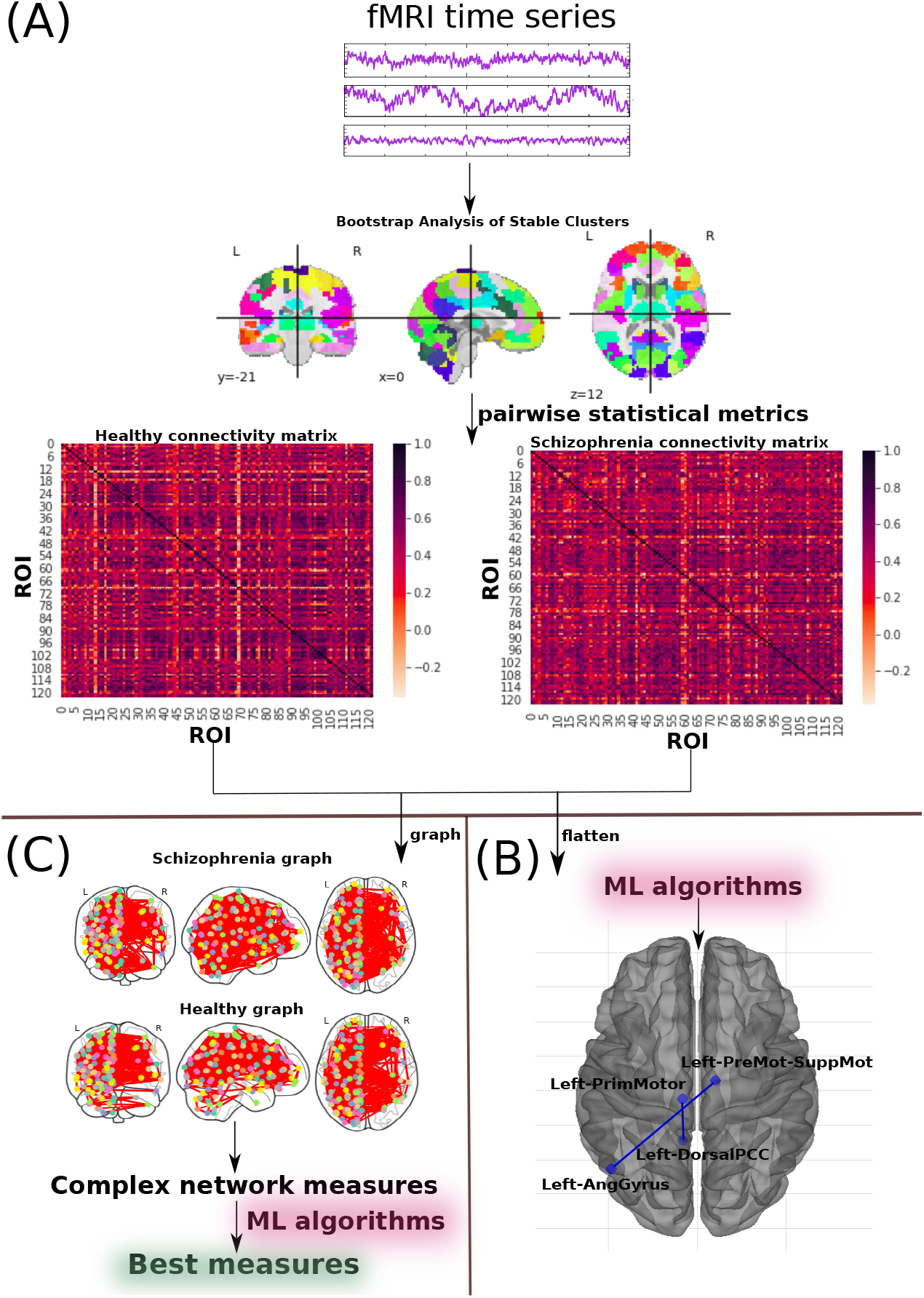
The methodology for diagnosing schizophrenia using fMRI schizophrenia data is also in subsection II A. (A) **fMRI preprocessing and selecting best pairwise metrics** methodology described in subsection II A 1; (B) **Connectivity matrix** methodology reported in subsection II A 2; (C) **Complex network measure** methodology described in subsection II A 3.

Further, Figure 2 fully represents the EEG entire method workflow used and organized into three parts accordingly to their aim. First, following preprocessing, the most distinguishing metrics discovered for the fMRI data were used to create a connection matrix (described in Figure 2-(A) and subsection II B 1), the best brain connection (described in Figure 2-(B) and in subsection II B 2), and the best complex network measures for differentiating schizophrenia from the control group (described in Figure 2-(C) and subsection II B 3.)

**FIG. 2:**
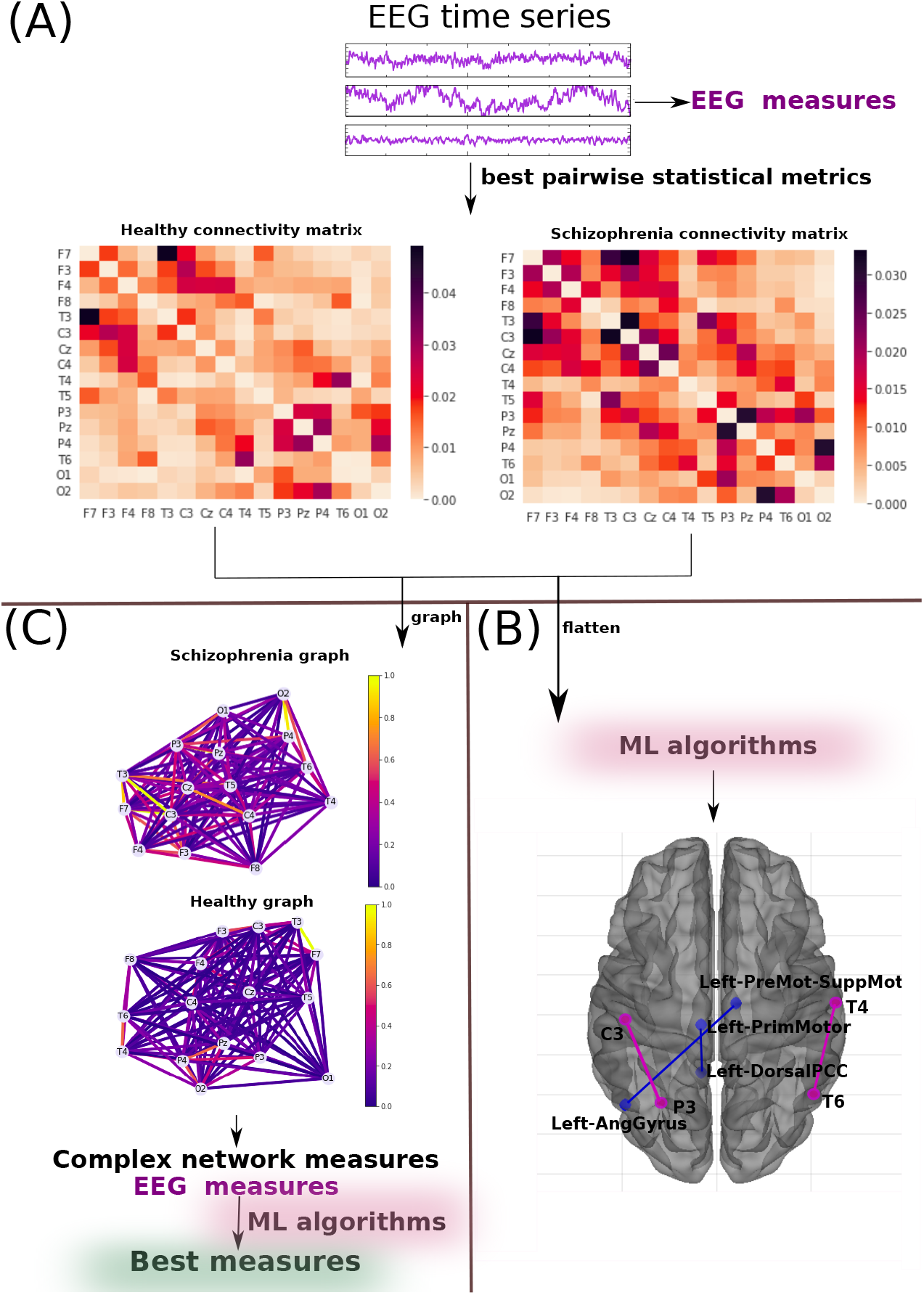
The methodology used here for the diagnosis of schizophrenia using EEG schizophrenia data in subsection II B. (A) **EEG preprocessing and selecting best pairwise metrics** methodology described in subsection II B 1; (B) **Connectivity matrix** methodology reported in subsection II B 2; (C) **Complex network measure** methodology described in subsection II B 3.

The python code with the methodology used in this work is available at: https://github.com/Carol180619/Paper-multimodal-schizophrenia.git.

### A. fMRI data

#### 1. fMRI data preprocessing and selecting best pairwise metrics

The fMRI data utilized in this study were from The Centers of Biomedical Research Excellence (COBRE) and included raw anatomical and fMRI data from 72 schizophrenia patients and 74 healthy controls (ages 18 to 65 in each group) with 6 minutes resting-state BOLD time series. As exclusion criteria, all the screened patients were excluded if they had a history of mental retardation, a neurological condition, severe head trauma with more than 5 minutes of loss of consciousness, or a history of drug dependency or misuse within the previous 12 months. More details can be seen in [38]. This data was accessed using the Nilearn Python library, in which the images were already preprocessed using the NIAK resting-state pipeline [39].

Brain regions of interest (ROIs) are considered rather than the whole BOLD time series collected from each voxel of the brain imaging. Because a brain atlas comprising these ROIs is employed, only the BOLD time series voxels of these ROIs were utilized. Bootstrap Analysis of Stable Clusters (BASC) was chosen from among the several preconfigured atlases since it was the map with the most outstanding performance according to [40, 41]. It was proposed in [42] and obtained via group brain parcellation using the BASC technique, a k-means clustering-based approach that finds brain networks with coherent activity in resting-state fMRI [43]. BASC map with a cluster number of 122 ROIs is used here (see Figure 1-(A)). Further, manual use of Yale BioImage Suite Package web application[44] labeled the coordinates of each ROIs for the identification of their names.

Once the time series for each of the 122 regions had been extracted, they were correlated according to Pearson Correlation (PC) [45], Spearman Correlation (SC) [46], Granger Causality (GC), [47], Biweight Midcorrelation (BM) [48], Sparce Canonical Correlation analysis (SCC)[49], Graphical Lasso method (GL) [50], Ledoit-Wolf shrinkage (LW) [51], Mutual Information (MI) [52], and Transfer Entropy (TE) [53] [54].

Each matrix was reduced to the size of the vectors used as input to the ML algorithm. The support vector machine (SVM) algorithm [55] was used as a classifier to select the most effective method to choose the best methods to construct the correlation and connectivity matrices. We use this method because it has been considered in studies of schizophrenia (see section I) and has a low computational cost, it also checked whether the use of metrics was better than the direct use of time series – the one of better performance would be chosen.

#### 2. Most important brain connection

After the best brain connectivity metric had been determined, the following ML classifiers were used: Random Forest (RF) [56], Naive Bayes (NB) [57], Multilayer Perceptron (MLP) [58], tuned Convolution Neural Network (called here *CNN*_*tuned*_ and *CNN*_*untuned*_) implemented in [59], and Long Short-Term Memory neural networks (LSTM) [60]. In addition to the CNN deep learning used in prior work [37], the LSTM network is a form of recurrent neural network commonly used to identify patterns in time series. Subsequently, the SHAP value method was used for the biological interpretation, as it explains the predictive power of each attribute. The same sampling data set was used in all ML algorithms and split into training (train) and test sets, with 25% data comprising the test set. was employed, with k = 10 which is a common value for this method [61–65]). This procedure is applied for model selection and hyper-parameter optimization. It was also considered the grid search method used for all ML algorithms except the untuned CNN and LSMT model, as done in [66–70]. The hyper-parameter optimization values for each classifier model are provided in Appendix B. The standard performance metric accuracy [71–75] was employed for evaluation. Due to the two-class (negative and positive) classification problem, other common metrics such as precision and recall were considered [76–79]. Precision (also called positive predictive value) corresponds to the hit rate in the negative class (here corresponding to the control group), whereas recall (also called sensitivity) measures how well a classifier can predict positive examples (hit rate in the positive class), here related to SCZ patients. Regarding the visualization of the two latter measures, the Receiver Operating Characteristic (ROC) curve is a common method that displays the relation between the rate of true and false positives. The area below the curve, called Area Under ROC Curve (AUC), has been widely used in classification problems [69, 71, 80, 81]. The AUC value ranges from 0 to 1-1 corresponds to a classification result free of errors, and 0.5 indicates the classifier cannot distinguish the classes, as in a random choice. The micro average of the ROC curve, which computes the AUC metric independently for each class (it calculates AUC for healthy individuals, class zero, and separately calculates it for unhealthy ones, class one), was also considered. The average is computed considering the classes equally. The macro average was also employed in our evaluation - it does not consider the classes equally but aggregates their contributions separately and then calculates the average.

#### 3. Best complex network measures

A complex network (or a graph) was generated for each connectivity matrix for the extraction of different measures. Towards inputting data into the ML algorithm, the complex network measures were stored in a matrix of attributes, where each column represents a complex network measure (feature), and each row denotes a subject. 2D matrices were generated for all subjects, as in [21].

To describe the network structure of the brain, the following complex network measures were calculated: assortativity coefficient [82, 83], average shortest path length (APL) [84], betweenness centrality (BC) [85], closeness centrality (CC) [86], eigenvector centrality (EC) [87], diameter [88], hub score [89], average degree of nearest neighbors [90] (Knn), mean degree [91], second moment of the degree distribution (SMD) [92], entropy of the degree distribution (ED) [93], transitivity [94, 95], complexity, k-core [96, 97], eccentricity [98], density [99], and efficiency [100].

Newly developed metrics (described in detail in [21]) reflecting the number of communities in a complex network were also applied. Community detection algorithms were also used in our study [101–103]. Since the community detection measures must be transformed into a single scalar value to be included in the matrix, community detection algorithms were applied to find the largest community. The average path length within the community was then calculated and received a single value as a result. The community detection algorithms used were the fastgreedy (FC) [104], Infomap (IC) [105], leading eigenvector (LC) [106], label propagation (LPC) [107], edge betweenness (EBC) [108], spinglass (SPC) [109], and multilevel community identification (MC) [110]. The abbreviations were extended with the letter “A” (for average path length) to indicate the approach (AFC, AIC, ALC, ALPC, AEBC, ASPC, and AMC).

These network measures were used to characterize the brain’s network structure. Thus, each observation representing the network properties of one patent is represented by a vector containing these metrics. The results are provided in subsection III A 2.

### B. EEG data

#### 1. Preprocessing and selecting best pairwise metrics

The EEG dataset used for diagnosis of SCZ, also used in [37], contains a 16-channel EEG time series recorded at a sampling frequency of 128 Hz over one minute, including *F*_3_, *F*_4_, *F*_7_, *F*_8_, *T*_3_, *C*_3_, *C*_*z*_, *C*_4_, *T*_4_, *T*_5_, *P*_3_, *P*_*z*_, *P*_4_, *T*_6_, *O*_1_, and *O*_2_. The study included 39 healthy young people (control group; aged 11 to 14 years) and 45 teenagers (aged 11 to 14 years) with schizophrenia symptoms in a resting state.

From these time series are extracted EEG measurements which are widely used in the literature, as there are spectral entropy [111, 112], Hjorth mobility and complexity [113–115] and Lempel-Ziv complexity [116, 117]. Further, connectivity matrices were generated with the most successful method evaluated for fMRI data; see section II A.

#### 2. Most important brain connection

Based on these connectivity matrices, the best ML method is used. With the SHAP value method, most distinguishing brain regions are found.

#### 3. Complex network measures

The same measures of complex networks used in the previous subsection were extracted. Moreover, for the first time in the literature, EEG measures extracted from time series and complex network measures have been put into the ML algorithm to obtain which metric best differentiates EEG data from schizophrenia patients.

## III. RESULTS

In general, ML algorithms were applied for two different levels of data abstraction, namely (B) the connectivity matrix and (C) the matrix of attributes, whose elements are complex network measures calculated from (B). This approach was used for EEG and fMRI data. However, for EEG, not only the complex network but also EEG measures extracted from time series.

We verified that all approaches automatically detected changes in the brain of SCZ patients. The fMRI connectivity matrix obtained the highest classification performance with a 99% mean AUC (see Table II and subsections III A and III B.

**TABLE II:**
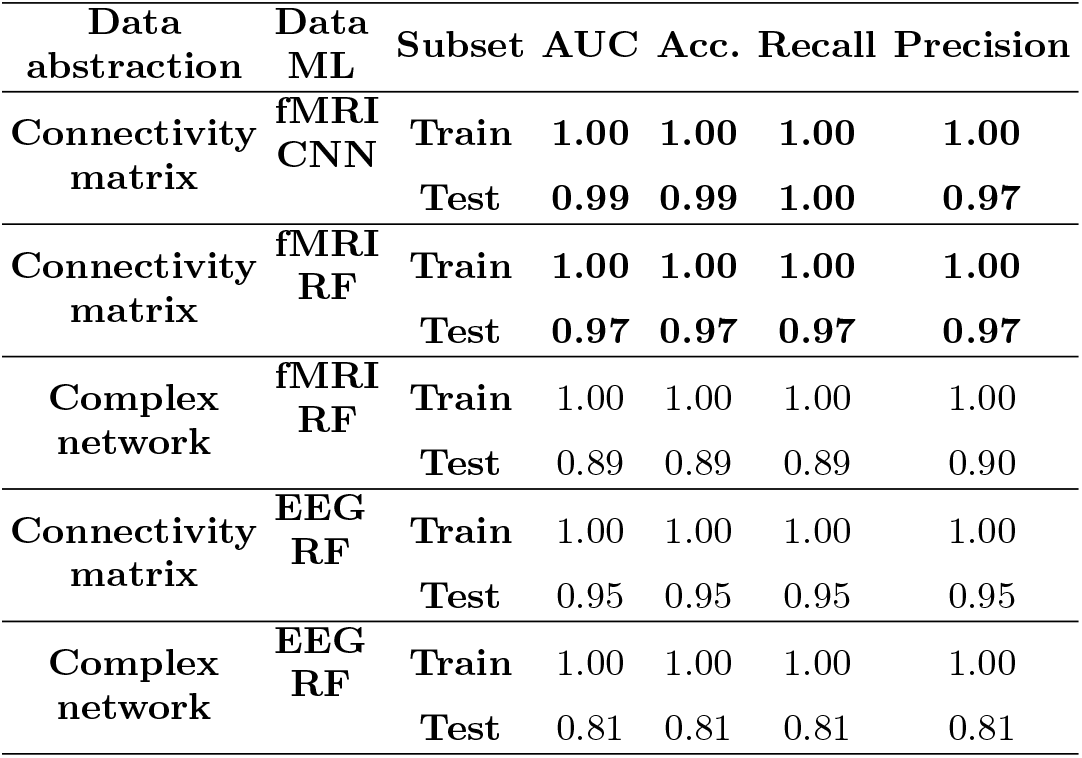
Summary of all the results obtained in the present work. Classification results using the connectivity matrix best-captured brain changes due to SCZ. The best performance is highlighted in bold.

**TABLE III:** Results from different ML algorithms. The best ML was *CNN*_*tuned*_ and RF for the fMRI dataset, whose performances are highlighted.

| ML methods | Subset | AUC | Acc. | Recall | Precision |
| --- | --- | --- | --- | --- | --- |
| RF | Train | **1.00** | **1.00** | **1.00** | **1.00** |
|  | Test | **0.97** | **0.97** | **0.97** | **0.97** |
| CNN tuned | Train | **1.00** | **1.00** | **1.00** | **1.00** |
|  | Test | **1.00** | **0.98** | **0.98** | **0.98** |
| CNN untuned | Train | 1.00 | 1.00 | 1.00 | 1.00 |
|  | Test | 1.00 | 0.98 | 1.00 | 0.97 |
| LSTM | Train | 1.00 | 0.99 | 0.99 | 0.99 |
|  | Test | 0.99 | 0.97 | 0.97 | 0.97 |
| SVM | Train | 0.99 | 0.99 | 1.00 | 0.99 |
|  | Test | 0.92 | 0.92 | 0.92 | 0.92 |
| NB | Train | 1.00 | 1.00 | 1.00 | 1.00 |
|  | Test | 0.87 | 0.86 | 0.88 | 0.89 |
| MLP | Train | 0.89 | 0.85 | 1.00 | 0.90 |
|  | Test | 0.80 | 0.80 | 0.80 | 0.80 |

### A. fMRI results

#### 1. Selecting best pairwise metrics

Appendix A contains the results for each connectivity matrix with different types of pairwise statistical metrics. SVM was used to detect the best one for capturing the brain changes due to SCZ in fMRI data. TE achieved the best performance. It is worth mentioning that the connectivity matrices outperformed the raw BOLD time series.

Then, the best connectivity matrix was tested with the other ML algorithms to determine the one that best-differentiated SCZ patients from control ones. According to Table III, the best classifiers are the *CNN*_*tuned*_ and RF. CNN performance for the test set was equal to 1.00 for the mean AUC and 0.98 for precision, recall, and accuracy. RF performance for the test set was equal to 0.97 for the mean AUC, precision, F1, recall, and accuracy. Figure 4 displays the confusion matrix (4-(a)), the learning curve (Figure 4-(b)), and the ROC curve (4- (c)), respectively, using TE and *CNN*_*tuned*_. The learning curve contains the loss error in each epoch. The loss error is computed on training and validation, and its interpretation is how well the model performs for these two sets. It is the total error committed for each example in these train, and test samples [118].

**FIG. 3:**
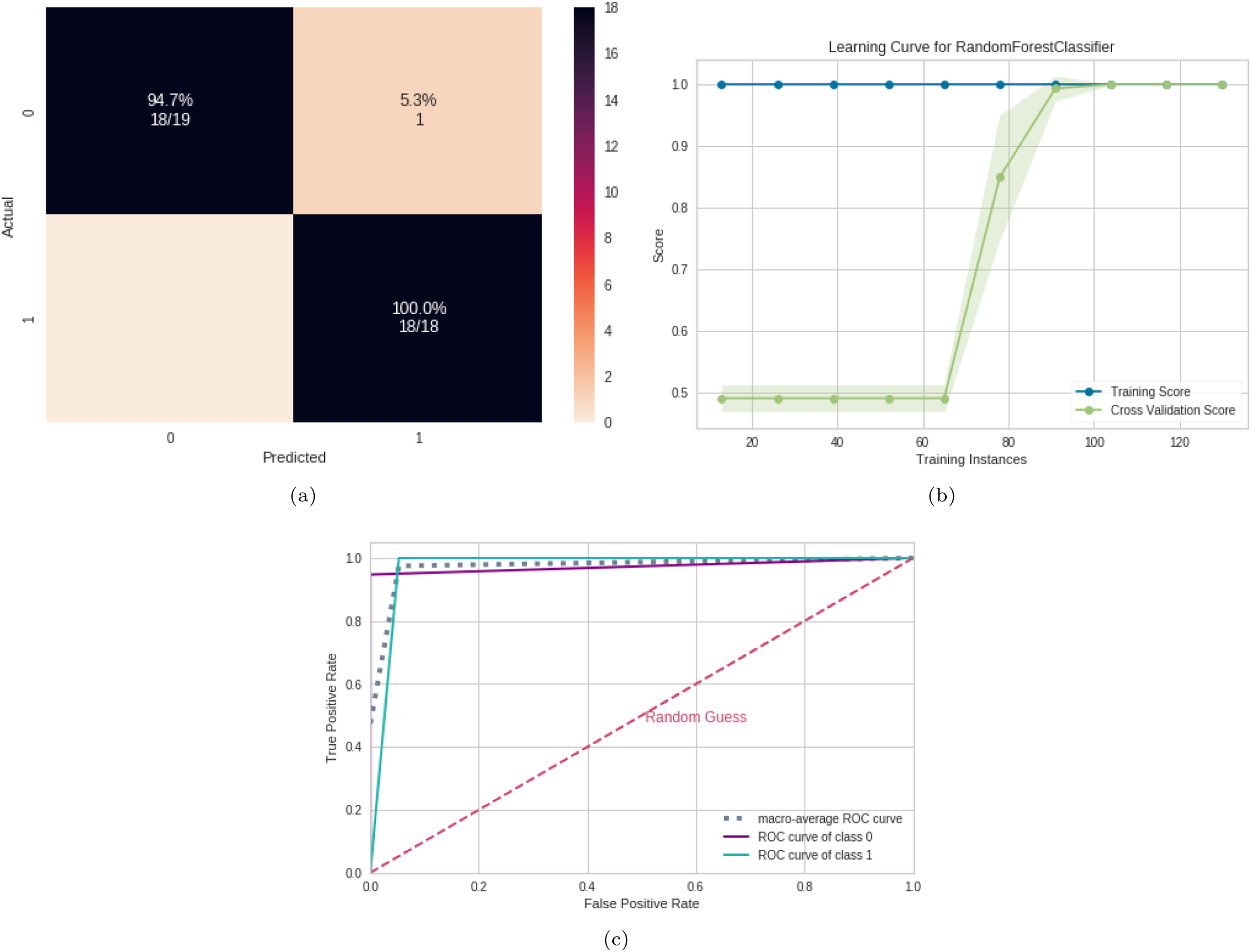
RF results using fMRI connectivity matrices. (a) Confusion matrix indicating a TN rate of 100% (purple, according to the color bar) and a TP rate of 95.2% (blue, according to the color bar). (b) Learning curve for the training Acc. (blue) and test Acc. (green). (c) ROC curve with class 0 (control) and class 1 (with SCZ).

**FIG. 4:**
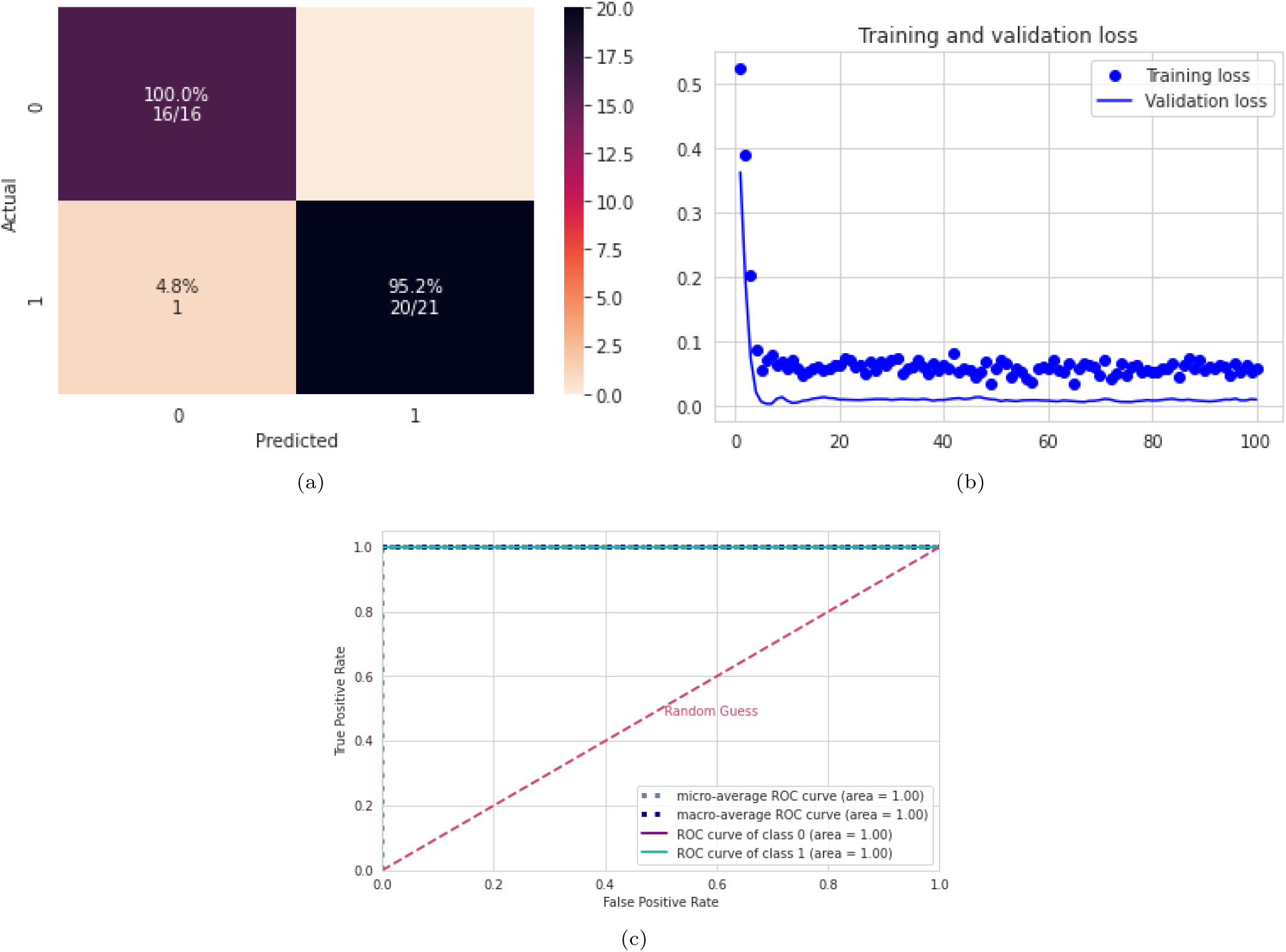
*CNN*_*tuned*_ results using fMRI connectivity matrices. (a) Confusion matrix indicating a TN rate of 100% (blue, according to the color bar) and a TP rate of 94.7% (blue, according to the color bar). (b) The learning curve with the Loss for the training (blue dots) and validation (line). (c) ROC curve with class 0 (control) and class 1 (with SCZ).

In contrast, Figure 3 displays the confusion matrix (3- (a)), the learning curve (Figure 3-(b)), and the ROC curve (3-(c)), respectively, using TE and RF. The results suggest that the complete database is not required to obtain the best validation accuracy (see learning curve in Figure 3- (b)). Regarding the classification model, TP (related to class 1) was higher than TN, showing that it better detects SCZ patients (see confusion matrix in Figure 3- (a)). The learning curve for ML assesses the model’s predictability by altering the size of the training set [30]. Since RF has a lower computational cost, it was chosen for the following steps.

SHAP values were calculated to quantify the importance of brain connections for the RF (see Figure 5 for the results). The connection between the Left-Dorsal Posterior Cingulate Cortex and Left-Primary motor cortex (Left-DorsalPCC – Left PrimMotor1) was the most important, according to Figure 5. Low correlation values (blue dots) for this connection (Left-DorsalPCC – Left PrimMotor1) were essential for the detection of the control group, and high values of this correlation (red dots) were important for the detection of SCZ. The second most crucial connection was detected between the Left-Premotor Cortex and Left-angular gyrus (Left-premotor suppmor5 – Left-AngGyrus1). Low correlation values for this connection (blue dots) were associated with SCZ patients, and high correlation values (red dots) were essential for detecting control ones. The corresponding brain regions are depicted in Figure 5.

**FIG. 5:**
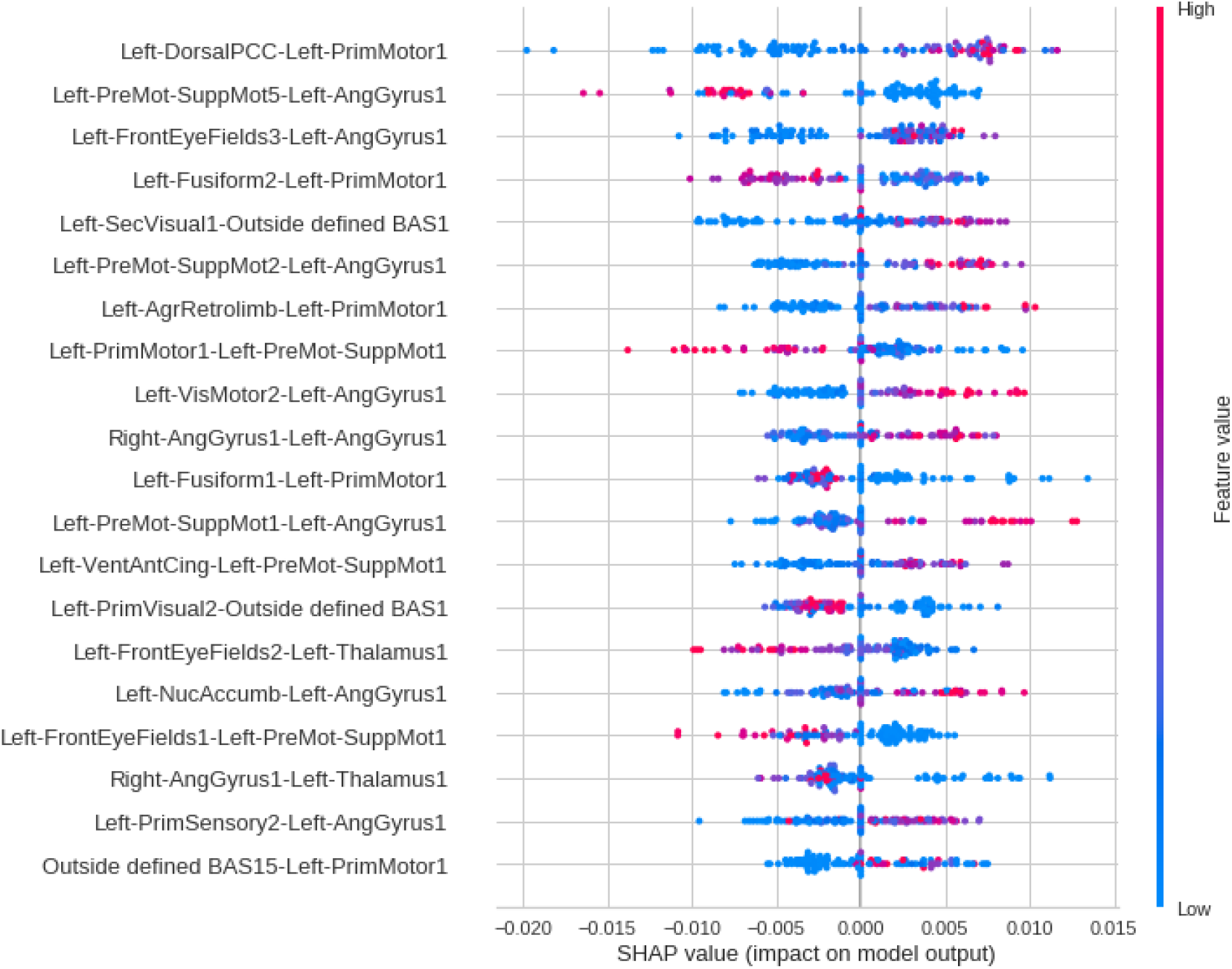
Feature importance ranking for the RF classifier with brain regions ranked in descending order of importance. For example, the connection between the regions Left-DorsalPCC and Left-PrimMotor1 is the most important to classify SCZ patients.

Since RF was the algorithm that provided the best performance, it was used in the following subsections.

#### 2. Complex network

The performance of the test sample considering the complex network yielded a mean AUC of 0.89, 0.90 for precision, 0.89 for F1 score, 0.89 for recall, and 0.89 for accuracy. Confusion matrix Figure 6-(a), learning curve Figure 6-(b), and ROC curve Figure 6-(c). Furthermore, according to Figure 6, the whole dataset was necessary. The results suggested that almost the complete database is required to obtain the best validation accuracy (see learning curve in Figure 6- (b)).

**FIG. 6:**
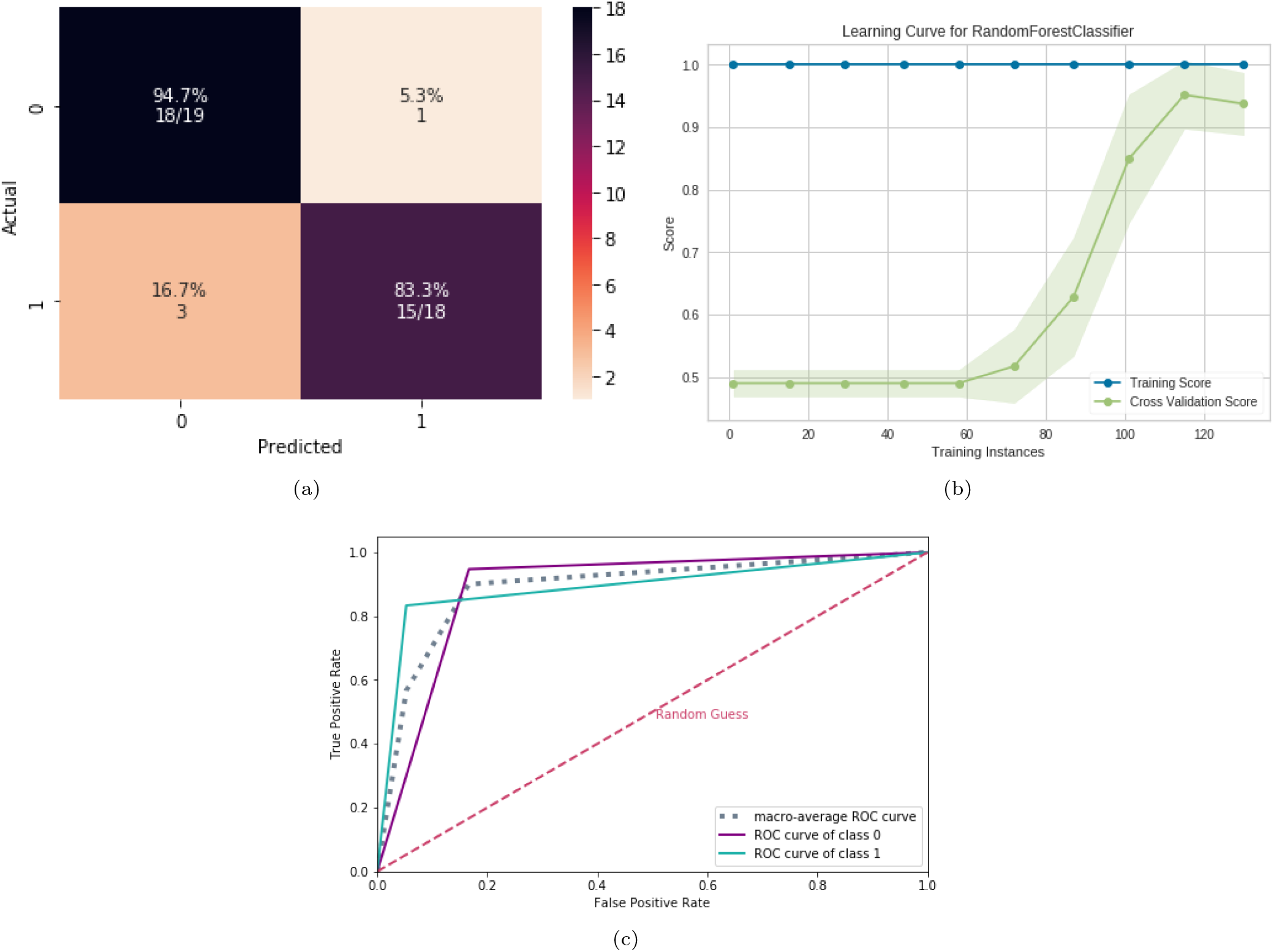
ML results using complex networks and fMRI. (a) Confusion matrix indicating a TN rate of (blue, according to the color bar) and a TP rate of (blue, according to the color bar). (b) Learning curve for the training Acc. (blue) and test Acc. (green). (c) ROC curve with class 0 (control) and class 1 (with SCZ).

According to the SHAP values in Figure 7, the most crucial measure for the model was the Diameter, followed by the CC. Furthermore, except for CC, it is difficult to determine whether low or high values of these other measures are related to the presence or absence of schizophrenia from the Figure 7. However, in contrast to the other measures, it is evident that low CC values are associated with the presence of schizophrenia.

**FIG. 7:**
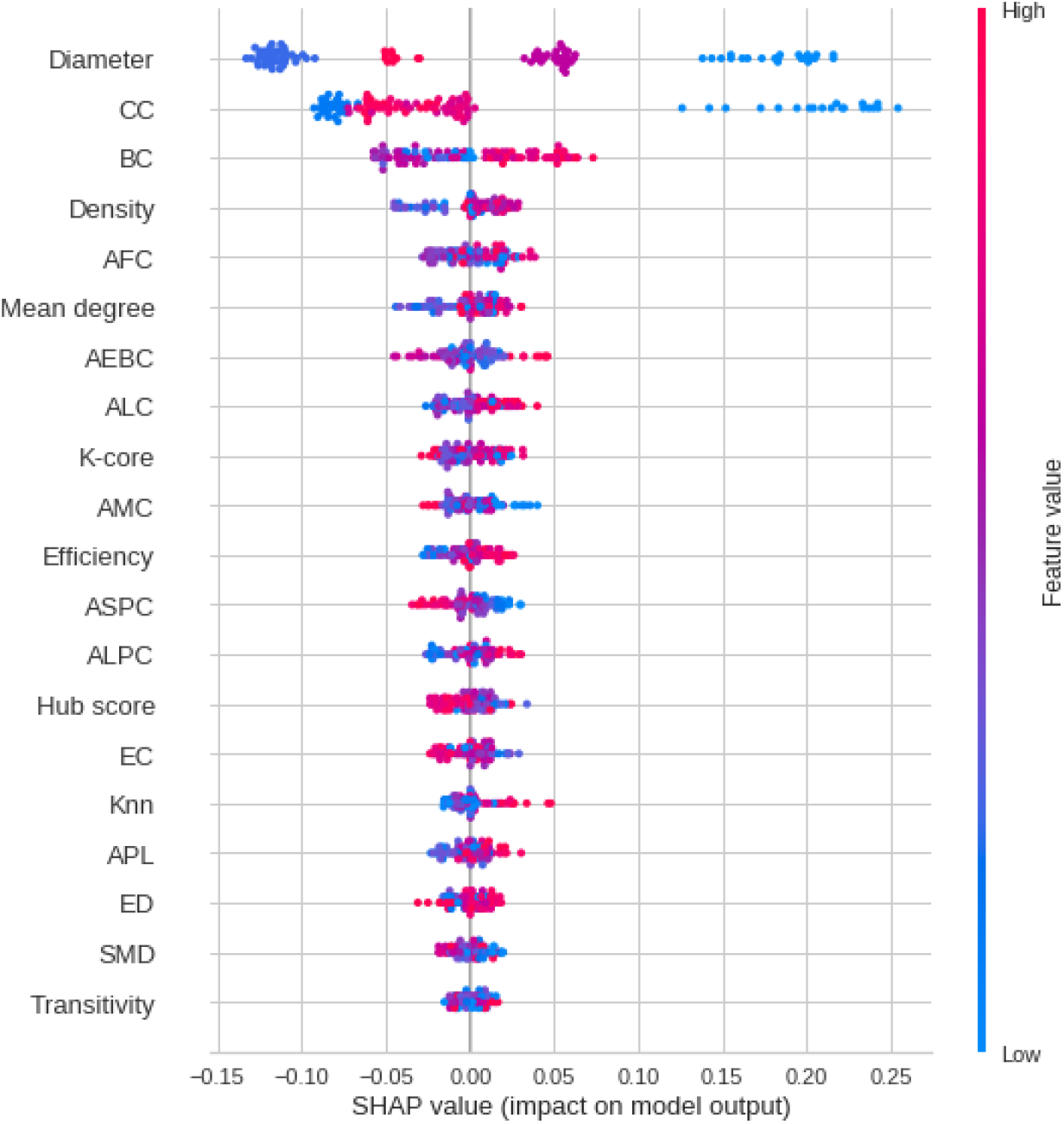
RF classifier features importance ranking in an fMRI dataset, with factors mentioned in descending order. For the categorization of SCZ patients, the spectral entropy measure is the most essential, followed by the Hjorth mobility and complexity measure.

### B. EEG dataset

#### 1. Connectivity matrix

As previously stated, the identical approach used in the preceding section was evaluated on the EEG data. Furthermore, the TE measure was utilized to build the connectivity matrices of the EEG data, and RF was used to differentiate SCZ patients from the control group. Its performance for the test set was equal to 0.95 for the mean AUC, precision, F1, recall, and accuracy. Figure 8 displays the confusion matrix (8-(a)), the learning curve (Figure 8-(b)), and the ROC curve (8-(c)), respectively. The results suggest that the complete database is required to obtain the best validation accuracy (see learning curve in Figure 8- (b)).

**FIG. 8:**
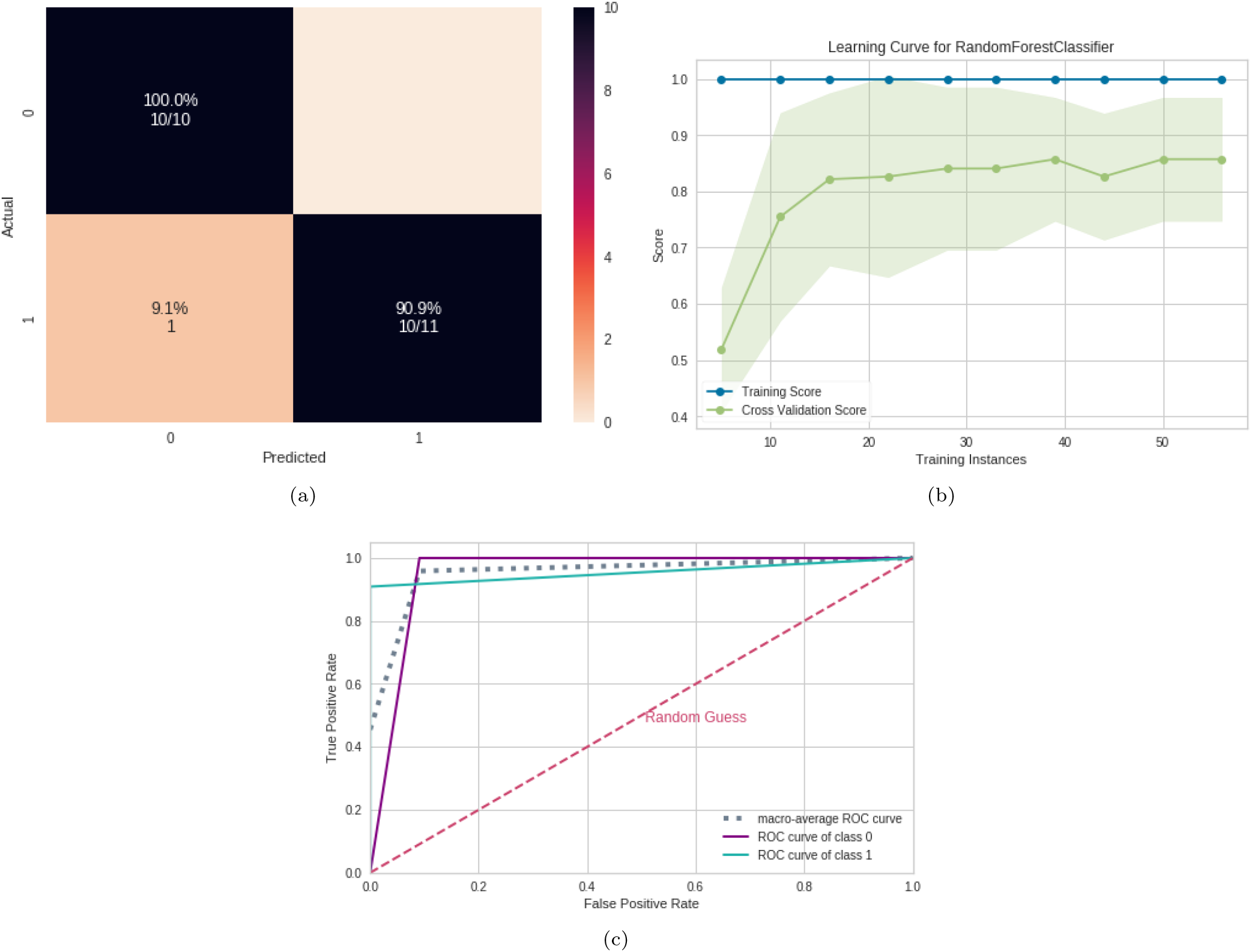
ML results using EEG connectivity matrices. (a) Confusion matrix indicating a TN rate of (blue, according to the color bar) and a TP rate of (blue, according to the color bar). (b) Learning curve for the training Acc. (blue) and test Acc. (green). (c) ROC curve with class 0 (control) and class 1 (with SCZ).

SHAP values were calculated to quantify the importance of brain connections for the RF (see Figure 9 for the results). The connection between regions P3 and C3 (P3 – C3) was the most important for the RF model. According to the data in Figure 9, high correlation values (red dots) for the connection (P3 – C3) were essential for the detection of SCZ patients, and low values of correlation (blue dots) were necessary for the detection of control ones. The second most crucial connection was detected between T6 and T4 (T6 – T3), and this connection’s low values were associated with SCZ. The corresponding brain regions are depicted in Figure 9

**FIG. 9:**
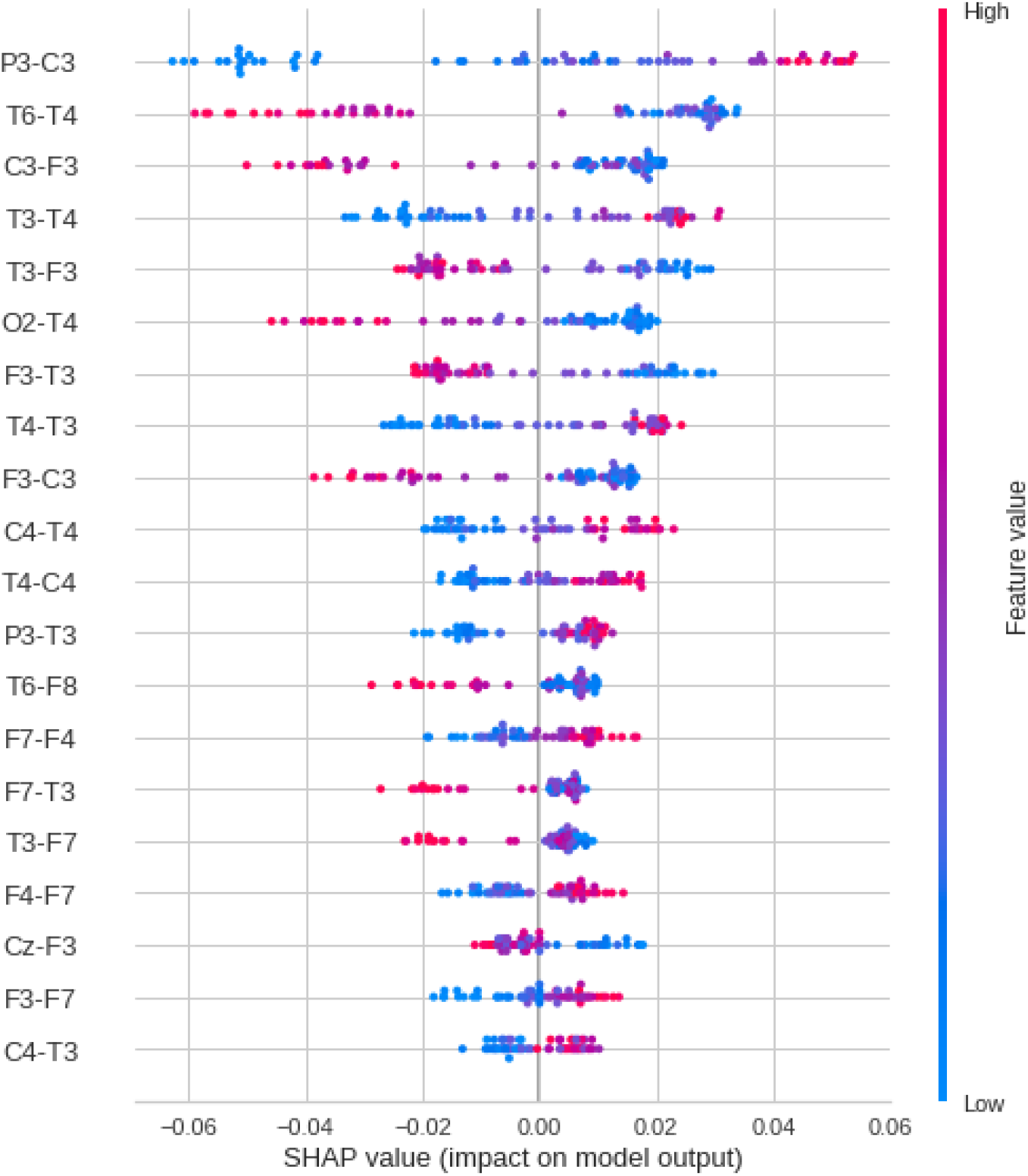
Feature importance ranking for the RF classifier with brain regions in descending order. The connection between the P3 and C3 is the most important for classifying SCZ patients.

#### 2. Complex network

The performance of the test sample considering the complex network yielded a mean AUC equal to 0.82, 0.85 for precision, 0.81 for F1 score, 0.75 for recall, and 0.81 for accuracy. Confusion matrix, learning curve, and ROC curve are shown in Figure 10. The results suggest that the complete database is required to obtain the best validation accuracy (see learning curve in Figure 10-(b)). Regarding the classification model, TP (related to class 1) was slightly higher than TN, showing that it better detects SCZ patients (see confusion matrix in Figure 10-(a)).

**FIG. 10:**
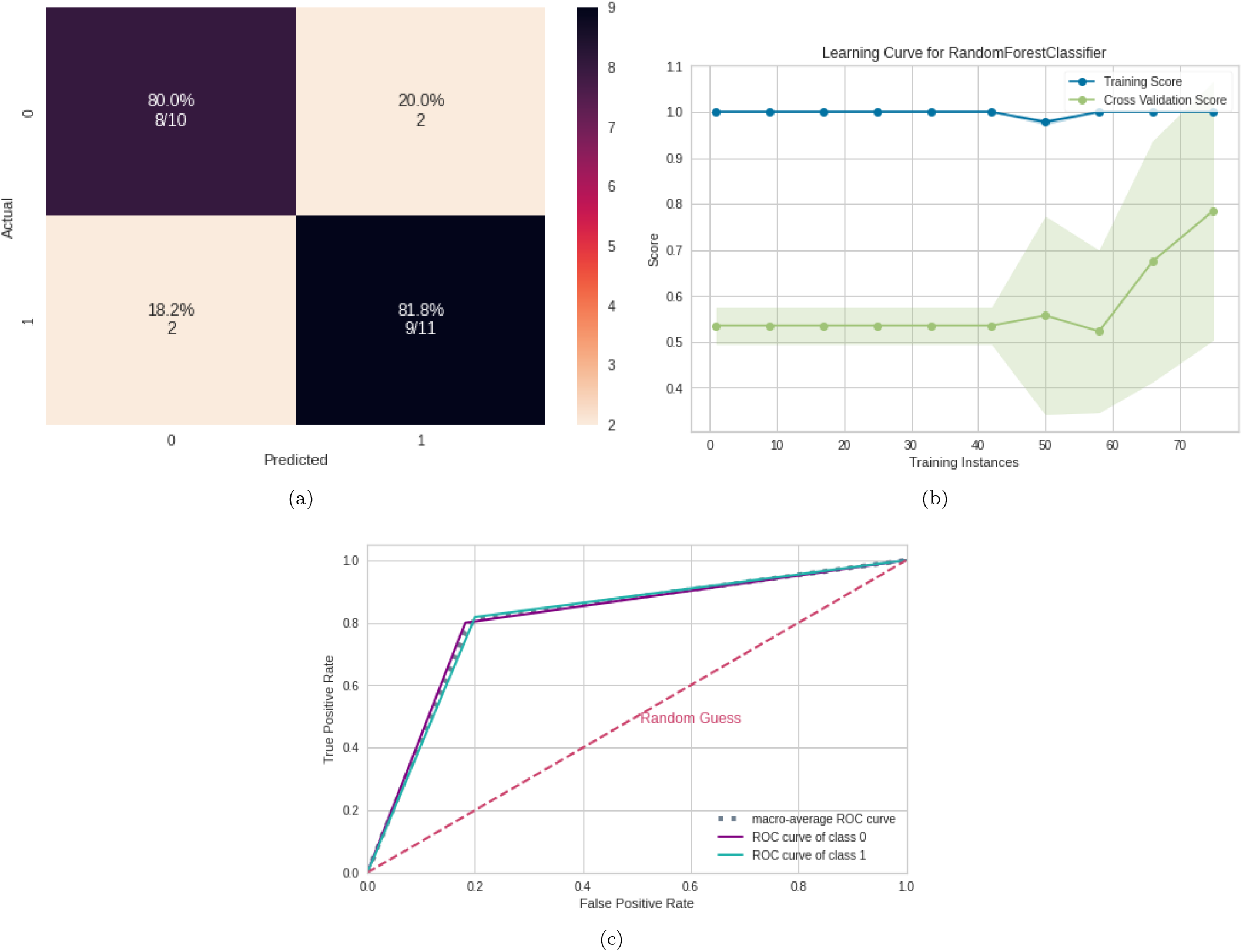
ML results using complex network measures from EEG. (a) Confusion matrix indicating a TN rate of (blue, according to the color bar) and a TP rate of (blue, according to the color bar). (b) Learning curve for the training Acc. (blue) and test Acc. (green). (c) ROC curve with class 0 (control) and class 1 (with SCZ).

According to the SHAP values in Figure 11, the most relevant measure was spectral entropy, which had a positive correlation with SCZ, followed by Hjorth mobility and complexity, which have a negative correlation with the occurrence of SCZ. Further, the measures extracted from the EEG time series were more important than the complex network measures. However, the diameter was the most relevant complex network measure, comparable to that found for the fMRI data. Furthermore, the existence of SCZ was connected with greater ASPC values, which estimated the size of the communities. As a result, SCZ is connected with larger communities.

**FIG. 11:**
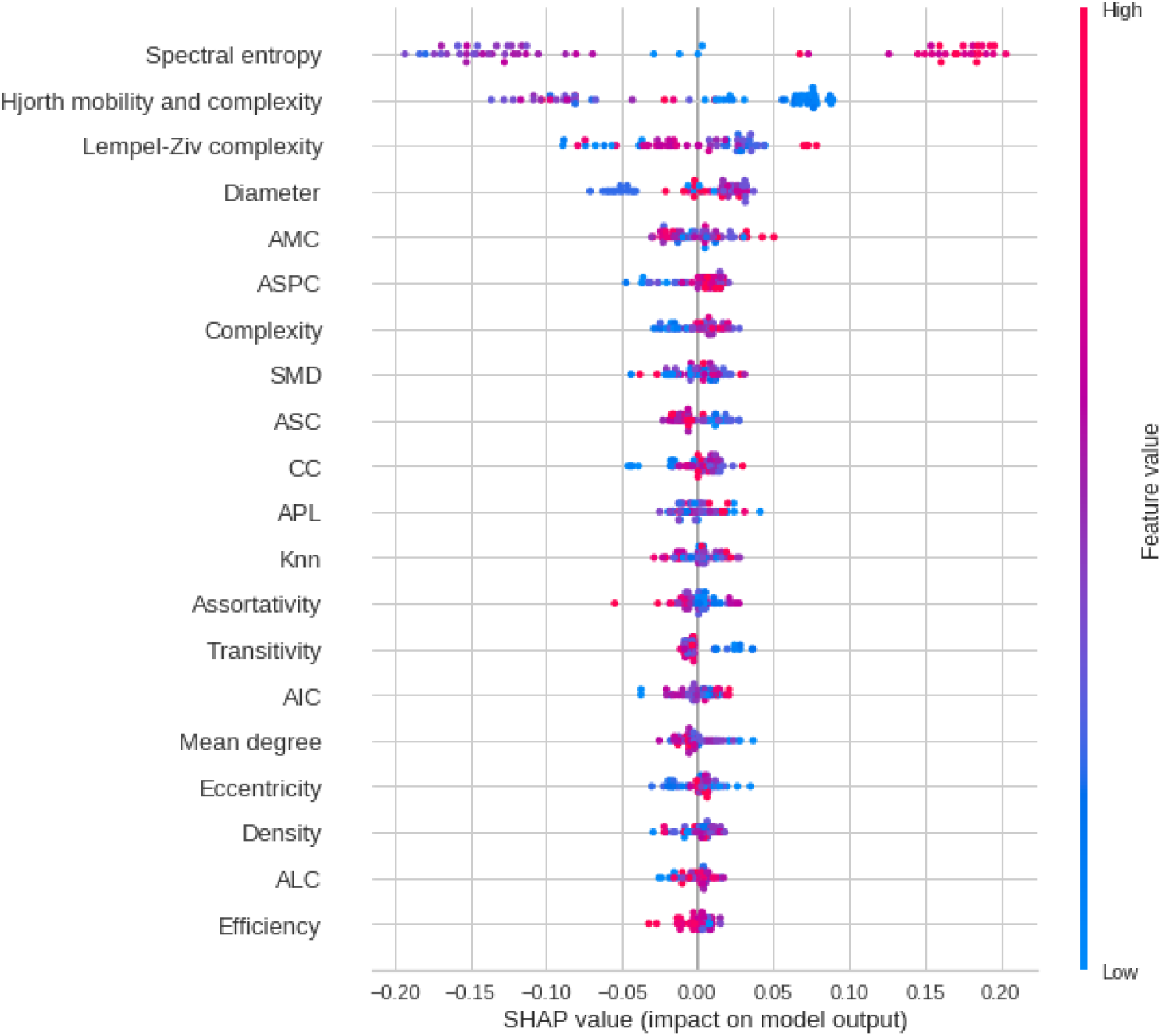
RF classifier features importance ranking in an EEG dataset, with factors mentioned in descending order. For the categorization of SCZ patients, the spectral entropy measure is the most essential, followed by the Hjorth mobility and complexity measure.

## IV. DISCUSSION

### A. Connectivity matrix

One aim of this work was to compare the classificatory statements that can be made based on two different SCZ datasets, namely fMRI and EEG. The ML workflow used was essentially the same. A comparison of the main results regarding altered connectivity in the neural network structure of the SCZ patients can be found in Figure 12.

**FIG. 12:**
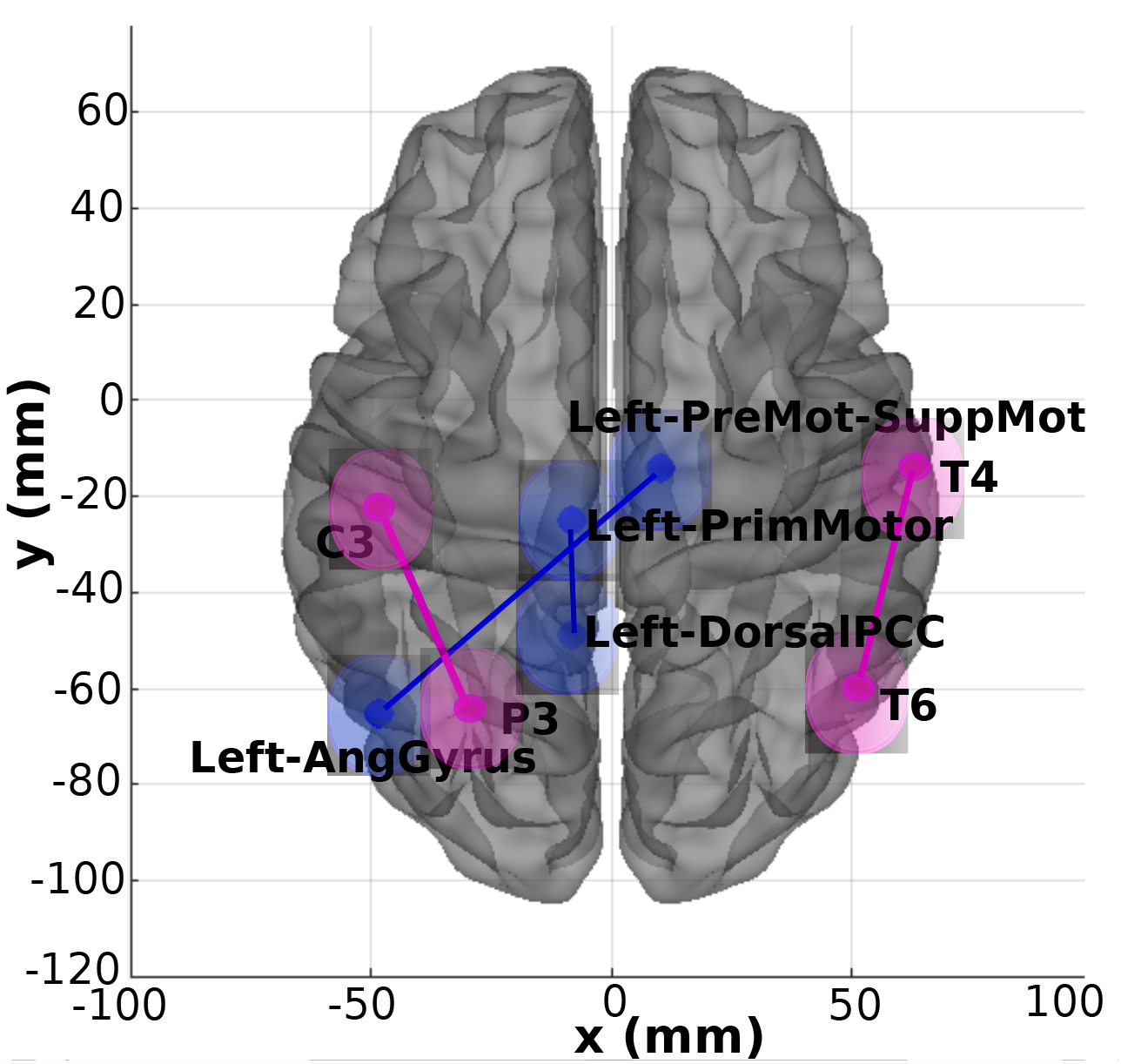
Plot with the most crucial connections found, in the two-dimensional schematic (ventral-axis), with the essential fMRI connection highlighted in blue and the most critical EEG connection highlighted in pink. The brain plot was developed by Braph tool [119], based on the coordinates in [120–122], and each region was plotted using the Brodmann map from the Yale BioImage Suite Package.

Our method identified a positive correlation in the SCZ group between the left dorsal PCC and the left primary motor during rest. The posterior cingulate cortex is in the upper region of the limbic system and is associated with Brodmann Areas (BA) 23 and 31. Researchers [123] suggested that this brain region behaves as a central hub for information exchange and has high connectivity with frontoparietal regions, which is related to the cognitive control of directing attention inside and outside. Furthermore, they observed that the ventral PCC is highly involved in DMN when there is the activation of cognitive activity directed towards the inside focus and when there is recovery and memory planning. On the other hand, dorsal PCC is related to highly complex connectivity directed to the frontal lobe, correlated to the balance of internal/external and broad/ narrow attention [123, 124].

In contrast, the primary motor cortex (PMC), BA04, was a brain region positively correlated with PCC in our study, usually activated when the finger is in movement [125] or when healthy subjects press a button in a given task. However, [126] suggested that the premotor and primary premotor cortex may be involved in language processing, especially the left premotor cortex, which performs articulation planning [127]. Considering previously cited studies mentioned here and in the I, the high positive correlation found between PCC and left PMC in our study suggests that activation of DMN-related areas, usually present during rest and reflecting voluntary targeting for inside observation, are also associated with language processing and may reflect problems in production, articulation, and speech expression, such as alterations in thought process manifested in speech, seen by [128] in SCZ patients.

The second highest correlation found in our study occurred between the left premotor supplementary (BA6) and angular gyrus (BA39) areas; this time, these areas are negatively correlated. The Supplementary Motor Area (SMA) has been associated with movement control and preparation [129], and patients with left medial SMA lesions showed severe difficulties remembering and reproducing rhythms compared to control subjects. Furthermore, it has been observed that bilateral SMA regions are altered in SCZ patients with catatonia, and this hyperperfusion is a marker of current catatonia in schizophrenia, indicating a dysregulation in the motor system, particularly affecting the premotor areas [130].

The angular gyrus brain region, located in the posterior inferior parietal area, is activated during different tasks, and as shown by [131] as being a hub that receives information and integrates it, generating comprehension and reasoning, redirecting attention to relevant information, manipulating mental processes, and problem-solving. Considering the angular gyrus findings described in the I and through the findings obtained for the negative correlation in our study, in the SCZ group compared to the control, between the left premotor supplementary regions and the angular gyrus, we can suggest that this correlation may reflect an altered motor system, with problems related to motor learning with altered co-ordination; express the cognitive deficits, language and thinking problems, both associated with SCZ.

We also observed a high positive correlation between P3 and C3 regions for the SCZ group compared to the control group. The P3 region corresponds to the Left Superior Parietal lobe (LSP), BA07, while the C3 region corresponds to the left motor cortex, BA06 [122]. LSP is activated during the performance of body part localization task [132]; both right and left parietal regions showed deficits in tests related to working memory information involving mental information [133]. The motor cortex is usually associated with motor actions. However, [134] highlighted its involvement in cognitive processes such as task-directed attention, motor consolidation, integration of multiple sensory inputs, and inhibition of involuntary movements. Therefore, the studies cited in I and the correlation observed between P3 and C3 regions in our study during resting state may reflect the dysfunctions found in SCZ that unfold working memory problems and deficits in motor cognition.

Our study’s negative correlation in the right temporal lobe between T6 and T4 electrodes for SCZ patients comprises BA 21 and 38. For example, the temporal lobe is associated with speech perception and production, hearing, and episodic memory [135]. During hallucinatory crises in SCZ patients, [136] found increased coherence in the temporal cortices bilaterally, suggesting abnormally increased synchrony in the left and right auditory cortices compared to the time without hallucination. In contrast with this study, we only found a correlation between two electrodes in the right temporal cortex in SCZ patients, indicating an inverse activity variation in this region. Furthermore, the hallucinating patients showed reduced alpha coherence in FT7, and FT8 electrodes, compared to the HC and the group of patients without hallucination [137]. This finding points to the importance of the activity of the temporal region bilaterally with other areas of SCZ patients.

### B. Complex network and measures extracted from EEG time series

The most important measure found for both data was Diameter, which corresponds to the length of the longest of the shortest path between any two vertices [138]. Since this measure was observed in two different bases, EEG and fMRI, it may be an indicative biomarker for the diagnosis of schizophrenia.

Regarding the EEG database, for the first time in the literature, we combined measures extracted from EEG time series with measures of complex networks extracted from functional networks of control individuals and schizophrenia patients to verify which measures are more efficient for this type of data. The results suggested that measures extracted from time series were more critical for classifying patients with SCZ. The most relevant measure for EEG was spectral entropy, which has a positive correlation with SCZ, followed by Hjorth mobility and complexity, which have a negative correlation with the occurrence of SCZ.

Spectral Entropy is an information theory-derived quantity that measures the degree of uncertainty in a signal, with higher values corresponding to a more uniform spectrum and more random frequency content and lower values corresponding to more regular frequencies [139, 140]. Spectral Entropy is an information theory-derived quantity that measures the degree of uncertainty in a signal, with higher values corresponding to a more uniform spectrum and more random frequency content and lower values corresponding to more regular frequencies. During the resting condition, we detected greater levels of spectral Entropy associated with SCZ, indicating more random frequencies. This conclusion contradicts previous studies [140–142] that reported spectral entropy deficiencies in schizophrenia patients executing the P300 activity. We assume this discrepancy because our data were obtained during the rest. The second most important EEG measure was Hjorth mobility and complexity, which illustrates a frequency shift and shows how a signal’s form is comparable to a pure sine wave [143]. The Hjorth mobility and complexity values found for SCZ patients were according to the literature [144].

Furthermore, for EEG data, the existence of SCZ was connected with greater ASPC values, which estimate the size of the communities. As a result, SCZ is connected with larger communities. Moreover, the discovery of larger communities shows that the brain’s equilibrium between functional segregation and integration has been disrupted, implying that information distribution is slower than in the control group. These findings support the idea that functional brain networks in SCZ patients are more random than the control ones, as described in section I.

Further, lower Transitivity values are related to the existence of SCZ. The transitivity is a measure of the efficiency of information transfer between all pairs of nodes in the graph [145], and a lower value of these measures indicates lower segregation [146]. This finding is according to the literature [33, 147].

Additionally, for fMRI, CC was the second most crucial measure. The average of the shortest path lengths from the node to every other node in the network, measured as CC, represents how near a node is to all other nodes in the network [148]. Therefore, a lower Closeness Centrality score reflects impairment at these nodes [149], which was also found in previous findings [150].

## V. CONCLUSIONS AND FUTURE WORK

The workflow developed using fMRI data distinguish control from SCZ patients with an accuracy of 98% and AUC of 100%. The best pairwise statistical metric captured brain changes due to the presence of SCZ was TE, and the best-performing machine learning model was RF.

According to the TE and RF, essential brain connections in the SCZ group are a positive correlation between the left dorsal PCC and left primary motor at rest and a negative correlation between the left premotor supplementary and angular gyrus areas. Furthermore, in the right parietal lobe, there is a strong positive correlation between the P3 and C3 regions and a negative correlation between the T6 and T4 electrodes, according to the EEG connectivity matrix.

Due to the employment of two databases collected in different groups of SCZ patients using two different types of equipment that capture detailed information on brain activity, this study has particular difficulties in comprehending the associations revealed (fMRI and EEG). However, the findings provide a wealth of information about brain activity in SCZ patients, which is corroborated by clinical and neurophysiological findings in the literature. We hypothesize that fMRI and EEG correlations in the data point to brain regions involved in motor, cognitive, and sensory processing, internal attention targeting, DMN-related regions, and auditory and language processing. This finding may reflect changes in SCZ patients, such as problems with expression, production, and speech articulation, changes in thinking, internal attention targeting, cognition, and hallucinatory episodes.

Concerning the complex network measures, Diameter is recognized as a critical measure for both data, and it may be a suggestive biomarker for the diagnosis of SCZ because the same result was obtained with two different equipment and patient groups. This study could be a significant finding, as it reveals a robust biomarker that enables ML-based diagnosis of schizophrenia disease regardless of data modalities. Furthermore, for EEG data, SCZ functional networks, compared to the control group, have larger communities and lower. Moreover, SCZ functional networks exhibit larger communities and lower Transitivity for EEG data, indicating slower information distribution and less segregation than in the control group. According to the literature, these results mean that our approach with the SHAP value method could predict the primary SCZ-related connections and the best complex network measures.

Nevertheless, according to our ML approach, EEG measures extracted from raw time series are more important than complex network measurements in capturing brain alterations in SCZ patients.

Finally, future studies may involve the application of the methodology to other fMRI data from ADHD-200 Global Competition. It can also be adopted with EEG data from patients with other neurological disorders, for example, dystonia [151].

## Data Availability

All data produced in the present work are contained in the manuscript

## VI. ACKNOWLEDGEMENTS

T.G.L.O.T acknowledges FAPESB (grant number 307/2020 – Cota 2020; BOL0202/2020) for the financial support. A.M.P. is indebted to FAPESP (grant 2019/22277-0) for the financial support. F.A.R. is indebted to CNPq (grant 309266/20190) and FAPESP (grant 19/23293-0) for the financial for the financial support provided to this research. C.T. gratefully acknowledges financial support from the Zentrum für Wisschenschaftliche Services und Transfer (ZeWiS) Aschaffenburg, Germany.

## Appendix A fMRI best pairwise metrics

Table IV contains the all pairwise metrics results.

**TABLE IV:**
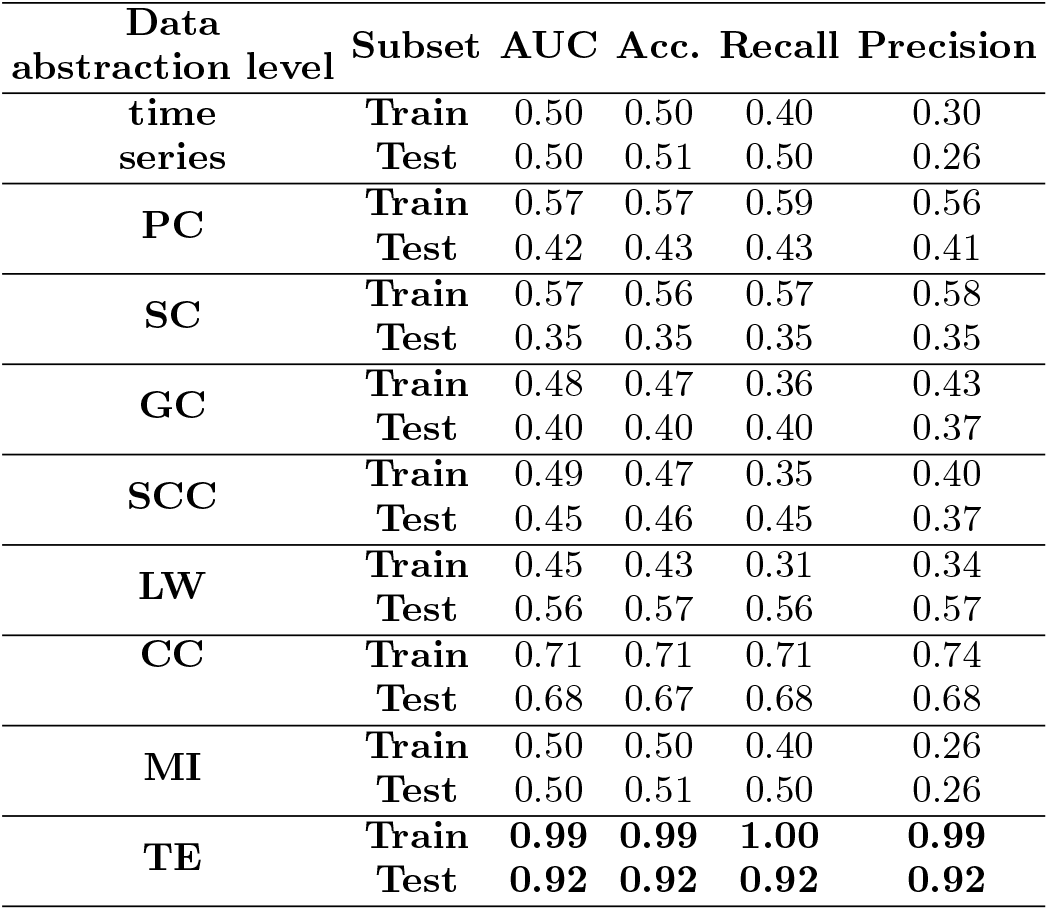
Results were obtained regarding the metrics used to obtain the connectivity matrix. The best metric was TE, whose performance is highlighted.

| Data abstraction level | Subset | AUC | Acc. | Recall | Precision |
| --- | --- | --- | --- | --- | --- |
| time series | Train | 0.50 | 0.50 | 0.40 | 0.30 |
|  | Test | 0.50 | 0.51 | 0.50 | 0.26 |
| PC | Train | 0.57 | 0.57 | 0.59 | 0.56 |
|  | Test | 0.42 | 0.43 | 0.43 | 0.41 |
| SC | Train | 0.57 | 0.56 | 0.57 | 0.58 |
|  | Test | 0.35 | 0.35 | 0.35 | 0.35 |
| GC | Train | 0.48 | 0.47 | 0.36 | 0.43 |
|  | Test | 0.40 | 0.40 | 0.40 | 0.37 |
| SCC | Train | 0.49 | 0.47 | 0.35 | 0.40 |
|  | Test | 0.45 | 0.46 | 0.45 | 0.37 |
| LW | Train | 0.45 | 0.43 | 0.31 | 0.34 |
|  | Test | 0.56 | 0.57 | 0.56 | 0.57 |
| CC | Train | 0.71 | 0.71 | 0.71 | 0.74 |
|  | Test | 0.68 | 0.67 | 0.68 | 0.68 |
| MI | Train | 0.50 | 0.50 | 0.40 | 0.26 |
|  | Test | 0.50 | 0.51 | 0.50 | 0.26 |
| TE | Train | **0.99** | **0.99** | **1.00** | **0.99** |
|  | Test | **0.92** | **0.92** | **0.92** | **0.92** |

## Appendix B Grid search hyperparameter tuning

Table V contains the hyperparameters optimized by gridsearch.

**TABLE V:** Hyperparameters for each classifier using Grid search optimizer.

| Classifier | Hyperparameters and description | Values |
| --- | --- | --- |
| **RF** | - max_depth: Maximum depth of the tree. | [1,2,5,10,20,80] |
|  | - max_features: Number of features to be considered toward a best split. | [2, 3,5,10] |
|  | - min_samples_leaf : Minimum number of samples required to be at a leaf node. | [1,2,3, 4, 5] |
|  | - min_samples_split: Minimum number of samples for the split of an internal node. | [1,2,8, 10, 12,20] |
|  | - n_estimators: Number of trees in the forest. | [1,2,3,5,10, 30,50,100, 200, 300,500] |
| **SVM** | -kernel: Specifies the kernel type to be used in the algorithm. | [rbf, linear] |
|  | -gamma: Kernel coefficient. | [1e-3, 1e-4] |
|  | -C: Regularization parameter. | [1, 10, 100, 1000] |
| **NB** | -var_smoothin:Portion of the largest variance of all features that is added to variances for calculation stability. | range 1e-09 to 1 |
| **MLP** | - activation: Activation function for the hidden layer. | [identity, logistic, tanh, relu] |
|  | - solver: Solver for weight optimization. | [lbfgs, sgd, adam] |
|  | - alpha: L2 penalty (regularization term) parameter. | [0.0001,1e-5,0.01,0.001] |
|  | - batch_size: Size of minibatches for stochastic optimizers. | [1000,5000] |
|  | - learning_rate: Learning rate schedule for weight updates. | [constant, invscaling, adaptive] |
|  | - learning_rate_init: The initial learning rate used. | [0.001,0.01,0.1,0.2,0.3] |
| **LR** | - C: Each value in Cs describes the inverse of regularization strength. | range 0.001 to 1000 |
|  | - penalty: Specifies the norm of the penalty. | [l1, l2] |

## Appendix C Deep Learning architetrure

Two approaches for the CNN architectures are proposed here, one using a Random Search tuning method (CNN_*tuned*_) and another without this optimization step (CNN_*untuned*_). Tuning is an optimization approach for determining hyperparameter values to improve the performance of the CNN model [152].

In the CNN_*tuned*_ model, the dropout regularization technique is employed to avoid overfitting [153]. The layers and range used for hyperparameters are presented in table VI. The best CNN_*tuned*_ architectures tuned for each data set are depicted in table VII. The CNN_*untuned*_ model presents fewer layers and, therefore, lower computational costs. The parameters used in our analysis are described in table VIII.

**TABLE VI:** Best hyperparameters and layer configurations obtained for the CNN_*tuned*_ model.

| Type of Layer | Tuning hyperparameter | Value |
| --- | --- | --- |
| Convolutional | — | — |
| Convolutional | dropout | [0.00, 0.05, 0.10, 0.15, 0.20, 0.25, 0.30, 0.35, 0.40, 0.45, 0.50] |
| Convolutional | — | — |
| Convolutional | number of filters | [32, 64] |
| Max Pooling | dropout | [0.00, 0.50, 0.10, 0.15, 0.20] |
| Flatten | — | — |
| Dense | - units | [32, 64, 96...512] |
|  | -activation | [relu, tanh, sigmoid] |
| Dropout | rate | [0.00, 0.50, 0.10, 0.15, 0.20] |
| Adam | | $min - value = 1e^{-4}$ |
| optimization | learning | $max - value = 1e^{-2}$ |
| compile | rate | sampling= LOG |

**TABLE VII:** The CNN_*tuned*_ model used in the SCZ dataset is the network architecture.

| Type of Layer | Output Shape (AD) | Parameter |
| --- | --- | --- |
| Convolutional | (None, 122, 122, 1) | 160 |
| Convolutional | (None, 119, 119, 32) | 2320 |
| max-pooling | (None, 59, 59, 32) | 0 |
| dropout | (None, 56, 56, 16) | 0 |
| Convolutional | (None, 28, 28, 16) | 4640 |
| Convolutional | (None, 2, 2, 32) | 9248 |
| max-pooling | (None, 1, 1, 32) | 0 |
| dropout | (None, 1, 1, 32) | 0 |
| flatten | (None, 32) | 0 |
| dense | (None, 160) | 5280 |
| dropout | (None, 160) | 0 |
| dense | (None, 2) | 3 |

**TABLE VIII:** The network architecture for the CNN_*untuned*_ model used in the SCZ dataset.

| Type of Layer | Output Layer | Kernel |
| --- | --- | --- |
| **Input Layer** | (None, 122, 122, 1) | 0 |
| **Convolution** | (None, 119, 119, 32) | 544 |
| **Max pooling** | (None, 59, 59, 32) | 0 |
| **Convolution** | (None, 56, 56, 16) | 8208 |
| **Max pooling** | (None, 28, 28, 16) | 0 |
| **Flatten** | (None, 12544) | 0 |
| **Fully connected** | (None, 10) | 125450 |
| **Fully connected** | (None, 1) | 11 |

The learning curve and ROC curve obtained for the CNN_*untuned*_ model are found in Figure 13.

**FIG. 13:**
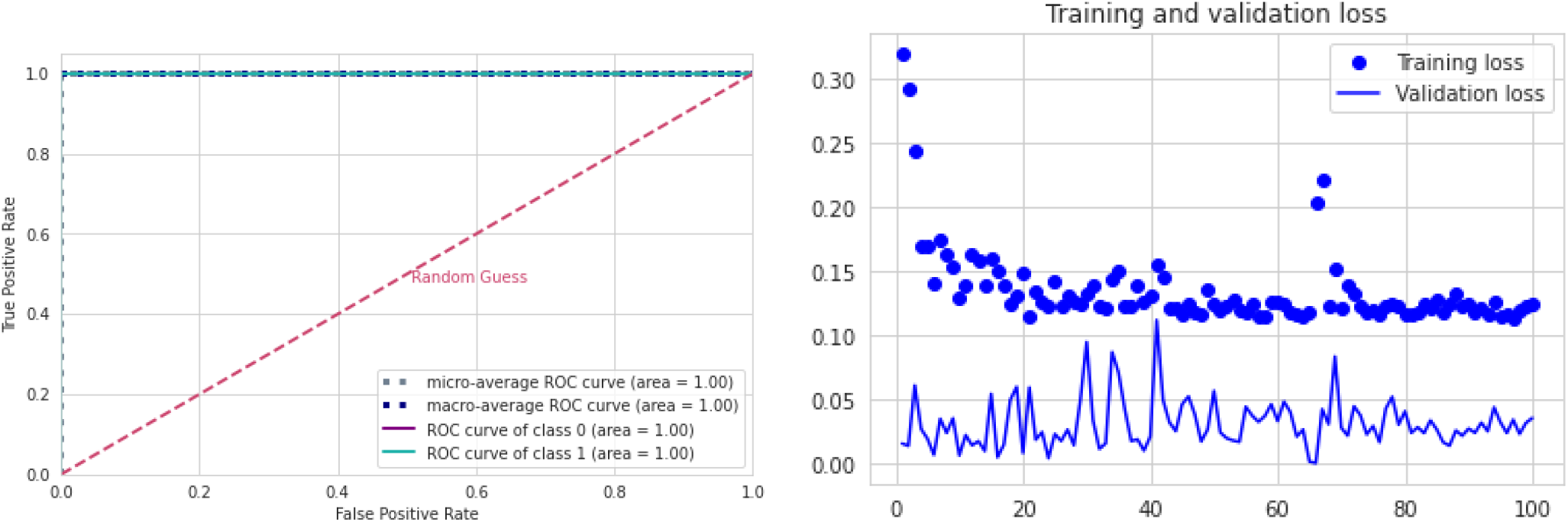
CNN_*untuned*_ model results using fMRI connectivity matrices. (a) ROC curve with class 0 (control) and class 1 (with SCZ). (b) Each epoch loses the training (blue dots) and validation (blue line).

Furthermore, the LSTM parameters used in our analysis are described in table IX.

**TABLE IX:**
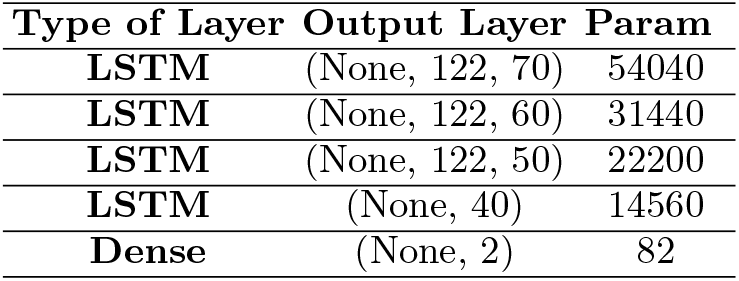
The network architecture for the LSTM model used in the SCZ dataset.

| Type of Layer | Output Layer | Param |
| --- | --- | --- |
| **LSTM** | (None, 122, 70) | 54040 |
| **LSTM** | (None, 122, 60) | 31440 |
| **LSTM** | (None, 122, 50) | 22200 |
| **LSTM** | (None, 40) | 14560 |
| **Dense** | (None, 2) | 82 |

The learning curve and ROC curve obtained for the LSTM model are found in Figure 14.

**FIG. 14:**
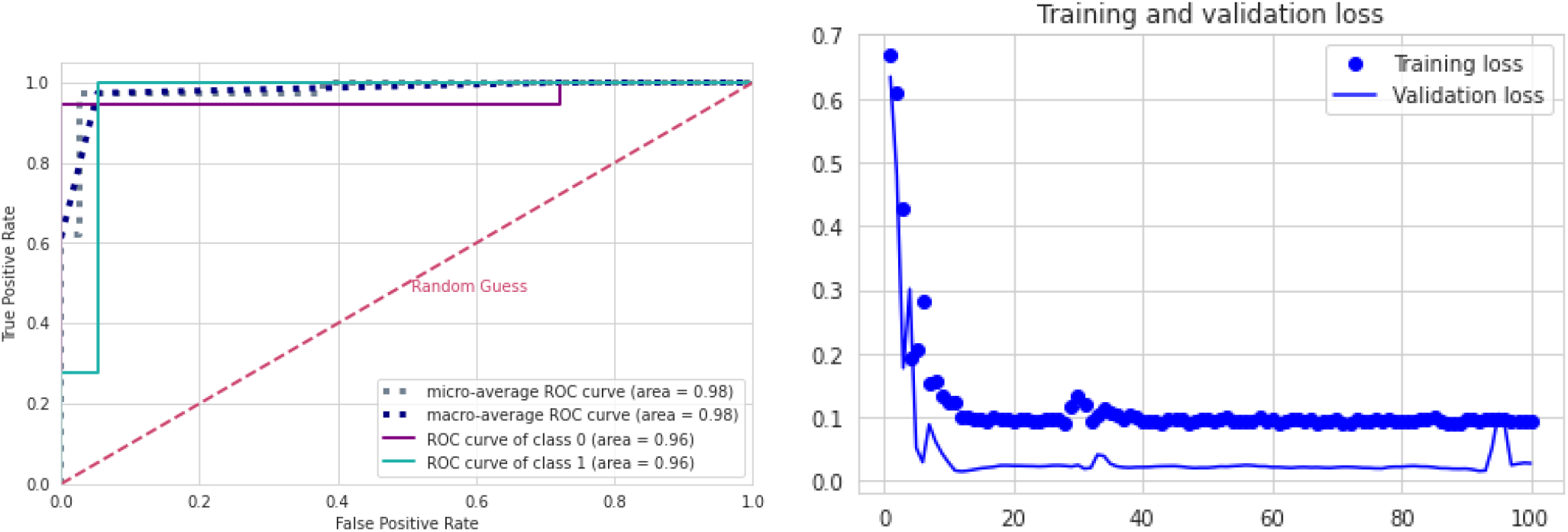
LSTM model results using fMRI connectivity matrices. a) ROC curve with class 0 (control) and class 1 (with SCZ). (b) Each epoch loses the training (blue dots) and validation (blue line).

## Appendix D Acronyms

### NOMENCLATURE

APL: Average shortest path length
AUC: Area Under ROC Curve
BA: Brodmann Areas
BASC: Bootstrap Analysis of Stable Clusters
BC: Betweenness centrality
BM: Biweight Midcorrelation
BOLD: Blood Oxygenation Level Dependent
CC: Closeness centrality
COBRE: The Centers of Biomedical Research Excellence
DMN: Default Mode Network
EBC: Edge betweenness community detection
EC: Eigenvector centrality
ED: Entropy of the degree distribution
EEG: Electroencephalogram
FC: Fastgreedy community detection
fMRI: Functional Magnetic Resonance imaging
GC: Granger Causality
GL: Graphical Lasso method
IC: Infomap community detection
Knn: Average degree of nearest neighbors
LC: Leading eigenvector community detection
LPC: Label propagation community detection
LSP: Left Superior Parietal Lobe
LSTM: Long short-term memory
LW: Ledoit-Wolf shrinkage
MC: Multilevel community detection
MI: Mutual Information
MLP: Multilayer Perceptron
NB: Naive Bayes
PC: Pearson Correlation
PCC: Posterior Cingulate Cortex
PMC: Primary Motor Cortex
RF: Random Forest
ROC: Receiver Operating Characteristic
ROIs: Regions of interest
SC: Spearman Correlation
SCC: Canonical Correlation analysis
SCZ: Schizophrenia
SHAP: SHapley Additive Explanations
SMA: Supplementary Motor Area
SMD: Second moment of the degree distribution
SPC: Spinglass community detection
TE: Transfer Entropy
Tuned: CNN tuned Convolution Neural Network
untuned: CNN Untuned Convolution Neural Network
WHO: World Health Organization

## References

[1] World Health Organization, https://www.who.int/news-room/fact-sheets/detail/schizophrenia (2021), [Online; Accessed on Septembre 21, 2021].

[2] H. B. Veague and C. E. Collins, Personality disorders (Infobase Publishing, 2007).

[3] E. Bleuler and C. G. Jung, Komplexe und krankheitsursachen bei dementia praecox, Zentralblatt fur Nerven-heilkunde und Psychiatrie, 220 (1908).

[4] O. Sporns, Networks of the Brain (MIT press, 2010).

[5] M. Huhn, A. Nikolakopoulou, J. Schneider-Thoma, M. Krause, M. Samara, N. Peter, T. Arndt, L. Bäckers, P. Rothe, A. Cipriani, et al., Comparative efficacy and tolerability of 32 oral antipsychotics for the acute treatment of adults with multi-episode schizophrenia: a systematic review and network meta-analysis, The Lancet 394, 939 (2019).

[6] N. C. Andreasen and M. Flaum, Schizophrenia: the characteristic symptoms, Schizophrenia bulletin 17, 27 (1991).

[7] Y. Bae, K. Kumarasamy, I. M. Ali, P. Korfiatis, Z. Akkus, and B. J. Erickson, Differences between schizophrenic and normal subjects using network properties from fmri, Journal of digital imaging 31, 252 (2018).

[8] K. J. Friston and C. D. Frith, Schizophrenia: a disconnection syndrome, Clinical Neuroscience 3, 89 (1995).

[9] V. D. Calhoun, T. Eichele, and G. Pearlson, Functional brain networks in schizophrenia: a review., Frontiers in human neuroscience (2009).

[10] H.-Y. Tan, S. Sust, J. W. Buckholtz, V. S. Mattay, A. Meyer-Lindenberg, M. F. Egan, D. R. Weinberger, and J. H. Callicott, Dysfunctional prefrontal regional specialization and compensation in schizophrenia, American Journal of Psychiatry 163, 1969 (2006).

[11] Y. Liu, M. Liang, Y. Zhou, Y. He, Y. Hao, M. Song, C. Yu, H. Liu, Z. Liu, and T. Jiang, Disrupted small-world networks in schizophrenia, Brain 131, 945 (2008).

[12] S. Micheloyannis, E. Pachou, C. J. Stam, M. Vourkas, S. Erimaki, and V. Tsirka, Using graph theoretical analysis of multi channel eeg to evaluate the neural efficiency hypothesis, Neuroscience letters 402, 273 (2006).

[13] M. Rubinov, S. A. Knock, C. J. Stam, S. Micheloyannis, A. W. Harris, L. M. Williams, and M. Breakspear, Small-world properties of nonlinear brain activity in schizophrenia, Human brain mapping 30, 403 (2009).

[14] Y.-j. Zhang, C.-c. Pu, Y.-m. Wang, R.-t. Zhang, X.-l. Cai, S.-z. Zhou, Y.-t. Ma, Y. Wang, E. F. Cheung, S. S. Lui, et al., Social brain network correlates with real-life social network in individuals with schizophrenia and social anhedonia, Schizophrenia Research 232, 77 (2021).

[15] A. Tyagi, V. P. Singh, and M. M. Gore, Towards artificial intelligence in mental health: a comprehensive survey on the detection of schizophrenia, Multimedia Tools and Applications,1 (2022).

[16] D. Dong, Y. Wang, X. Chang, C. Luo, and D. Yao, Dysfunction of large-scale brain networks in schizophrenia: a meta-analysis of resting-state functional connectivity, Schizophrenia bulletin 44, 168 (2018).

[17] J. Nierenberg, D. F. Salisbury, J. J. Levitt, E. A. David, R. W. McCarley, and M. E. Shenton, Reduced left angular gyrus volume in first-episode schizophrenia, American Journal of Psychiatry 162, 1539 (2005).

[18] M. Niznikiewicz, R. Donnino, R. W. McCarley, P. G. Nestor, D. V. Iosifescu, B. O’Donnell, J. Levitt, and M. E. Shenton, Abnormal angular gyrus asymmetry in schizophrenia, American Journal of Psychiatry 157, 428 (2000).

[19] M. Schürmann, J. Järveläinen, S. Avikainen, T. D. Cannon, J. Lönnqvist, M. Huttunen, and H. Riitta, Manifest disease and motor cortex reactivity in twins discordant for schizophrenia, The British Journal of Psychiatry 191, 178 (2007).

[20] F. De Vico Fallani, F. A. Rodrigues, L. da Fontoura Costa, L. Astolfi, F. Cincotti, D. Mattia, S. Salinari, and F. Babiloni, Multiple pathways analysis of brain functional networks from eeg signals: an application to real data, Brain topography 23, 344 (2011).

[21] C. L. Alves, R. G. Cury, K. Roster, A. M. Pineda, F. A. Rodrigues, C. Thielemann, and M. Ciba, Application of machine learning and complex network measures to an eeg dataset from ayahuasca experiments, medRxiv (2022).

[22] V. Menon and S. Crottaz-Herbette, Combined eeg and fmri studies of human brain function, Int Rev Neurobiol 66, 291 (2005).

[23] E. Formisano, D.-S. Kim, F. Di Salle, P.-F. Van de Moortele, K. Ugurbil, and R. Goebel, Mirror-symmetric tonotopic maps in human primary auditory cortex, Neuron 40, 859 (2003).

[24] The decrease in the rate of deoxyhemoglobin can be detected with the increase of the NMR signal. This effect is called Blood Oxygenation Level Dependent (BOLD).

[25] M. J. Sturzbecher, Detecção e caracterização da resposta hemodinâmica pelo desenvolvimento de novos métodos de processamento de imagens funcionais por ressonância magnética, Ph.D. thesis, Universidade de São Paulo (2006).

[26] B. Biswal, F. Zerrin Yetkin, V. M. Haughton, and J. S. Hyde, Functional connectivity in the motor cortex of resting human brain using echo-planar mri, Magnetic resonance in medicine 34, 537 (1995).

[27] E. d. O. Lopes et al., Análise de medidas em grafos para conectividade funcional em redes de modo padrão na demência da doença de alzheimer leve utilizando técnicas de aprendizado de maquina, (2016).

[28] D. Bowen and L. Ungar, Generalized shap: Generating multiple types of explanations in machine learning, arXiv preprint 2006.07155 (2020).

[29] R. Rodríguez-Pérez and J. Bajorath, Interpretation of compound activity predictions from complex machine learning models using local approximations and shapley values, Journal of Medicinal Chemistry 63, 8761 (2019).

[30] G. Spadon, A. C. de Carvalho, J. F. Rodrigues-Jr, and L. G. Alves, Reconstructing commuters network using machine learning and urban indicators, Scientific reports 9, 1 (2019).

[31] M. N. I. Qureshi, J. Oh, D. Cho, H. J. Jo, and B. Lee, Multimodal discrimination of schizophrenia using hybrid weighted feature concatenation of brain functional connectivity and anatomical features with an extreme learning machine, Frontiers in neuroinformatics 11, 59 (2017).

[32] P. Patel, P. Aggarwal, and A. Gupta, Classification of schizophrenia versus normal subjects using deep learning, in Proceedings of the Tenth Indian Conference on Computer Vision, Graphics and Image Processing (2016) pp. 1–6.

[33] A. Anderson and M. S. Cohen, Decreased small-world functional network connectivity and clustering across resting state networks in schizophrenia: an fmri classification tutorial, Frontiers in human neuroscience 7, 520 (2013).

[34] A. Savio and M. Granã, Local activity features for computer aided diagnosis of schizophrenia on resting-state fmri, Neurocomputing 164, 154 (2015).

[35] L.-L. Zeng, H. Wang, P. Hu, B. Yang, W. Pu, H. Shen, X. Chen, Z. Liu, H. Yin, Q. Tan, et al., Multi-site diagnostic classification of schizophrenia using discriminant deep learning with functional connectivity mri, EBioMedicine 30, 74 (2018).

[36] M. Al-Beltagi, Autism medical comorbidities, World journal of clinical pediatrics 10, 15 (2021).

[37] C. L. Alves, A. M. Pineda, K. Roster, C. Thielemann, and F. A. Rodrigues, Eeg functional connectivity and deep learning for automatic diagnosis of brain disorders: Alzheimer’s disease and schizophrenia, Journal of Physics: Complexity 3, 025001 (2022).

[38] V. D. Calhoun, J. Sui, K. Kiehl, J. Turner, E. Allen, and G. Pearlson, Exploring the psychosis functional connectome: aberrant intrinsic networks in schizophrenia and bipolar disorder, Frontiers in psychiatry 2, 75 (2012).

[39] P. Bellec, Cobre preprocessed with niak 0.17-lightweight release, DOI 10, m9 (2016).

[40] F. Z. Subah, K. Deb, P. K. Dhar, and T. Koshiba, A deep learning approach to predict autism spectrum disorder using multisite resting-state fmri, Applied Sciences 11, 3636 (2021).

[41] C. Alves, G. d. O. Thaise, P. de Carvalho Aguiar, A. M. Pineda, K. Roster, C. Thielemann, J. A. M. Porto, and F. A. Rodrigues, Diagnosis of autism spectrum disorder based on functional brain networks and machine learning, (2022).

[42] P. Bellec, P. Rosa-Neto, O. C. Lyttelton, H. Benali, and A. C. Evans, Multi-level bootstrap analysis of stable clusters in resting-state fmri, Neuroimage 51, 1126 (2010).

[43] X. Yang, N. Zhang, and P. Schrader, A study of brain networks for autism spectrum disorder classification using resting-state functional connectivity, Machine Learning with Applications 8, 100290 (2022).

[44] Avaiable in https://bioimagesuiteweb.github.io/webapp/mni2tal.html.

[45] J. Benesty, J. Chen, Y. Huang, and I. Cohen, Pearson correlation coefficient, in Noise reduction in speech processing (Springer, 2009) pp. 1–4.

[46] D. Lubinski, Introduction to the special section on cognitive abilities: 100 years after spearman’s (1904)”‘general intelligence,’objectively determined and measured”., Journal of personality and social psychology 86, 96 (2004).

[47] C. W. Granger, Investigating causal relations by econometric models and cross-spectral methods, Econometrica: journal of the Econometric Society, 424 (1969).

[48] R. R. Wilcox, Introduction to robust estimation and hypothesis testing (Academic press, 2011).

[49] D. R. Hardoon and J. Shawe-Taylor, Sparse canonical correlation analysis, Machine Learning 83, 331 (2011).

[50] S. Sojoudi, Equivalence of graphical lasso and thresholding for sparse graphs, The Journal of Machine Learning Research 17, 3943 (2016).

[51] O. Ledoit and M. Wolf, Nonlinear shrinkage estimation of large-dimensional covariance matrices, The Annals of Statistics 40, 1024 (2012).

[52] A. Kraskov, H. Stögbauer, and P. Grassberger, Estimating mutual information, Physical review E 69, 066138 (2004).

[53] t. Schreiber, Measuring information transfer, Physical review letters 85, 461 (2000).

[54] For the TE, MI and GL metrics a Min-max normalization and then a thresholding process was performed, with a value of 0.5, since these measures deal best with binary values.

[55] L. Bottou and C.-J. Lin, Support vector machine solvers, Large scale kernel machines 3, 301 (2007).

[56] L. Breiman, Random forests, Machine learning 45, 5 (2001).

[57] N. Friedman, D. Geiger, and M. Goldszmidt, Bayesian network classifiers, Machine learning 29, 131 (1997).

[58] G. Hinton, D. Rumelhart, and R. Williams, Learning internal representations by error propagation, Parallel distributed processing 1, 318 (1986).

[59] C. L. Alves, Diagnóstico de doenças mentais baseado em mineração de dados e redes complexas, Ph.D. thesis, Universidade de São Paulo.

[60] S. Hochreiter and J. Schmidhuber, Long short-term memory, Neural computation 9, 1735 (1997).

[61] D. Berrar, Cross-validation. (2019).

[62] Y. Bengio and Y. Grandvalet, No unbiased estimator of the variance of k-fold cross-validation, Journal of machine learning research 5, 1089 (2004).

[63] A. A. Shah and Y. D. Khan, Identification of 4-carboxyglutamate residue sites based on position based statistical feature and multiple classification, Scientific Reports 10, 1 (2020).

[64] t. Kawamoto and Y. Kabashima, Cross-validation estimate of the number of clusters in a network, Scientific reports 7, 1 (2017).

[65] J. Chan, T. Rea, S. Gollakota, and J. E. Sunshine, Contactless cardiac arrest detection using smart devices, NPJ digital medicine 2, 1 (2019).

[66] M. Sato, K. Morimoto, S. Kajihara, R. Tateishi, S. Shiina, K. Koike, and Y. Yatomi, Machine-learning approach for the development of a novel predictive model for the diagnosis of hepatocellular carcinoma, Scientific reports 9, 1 (2019).

[67] Z. Zhong, X. Yuan, S. Liu, Y. Yang, and F. Liu, Machine learning prediction models for prognosis of critically ill patients after open-heart surgery, Scientific Reports 11, 1 (2021).

[68] F. Arcadu, F. Benmansour, A. Maunz, J. Willis, Z. Haskova, and M. Prunotto, Author correction: Deep learning algorithm predicts diabetic retinopathy progression in individual patients, NPJ digital medicine 3, 1 (2020).

[69] C. Krittanawong, H. U. H. Virk, A. Kumar, M. Aydar, Z. Wang, M. P. Stewart, and J. L. Halperin, Machine learning and deep learning to predict mortality in patients with spontaneous coronary artery dissection, Scientific reports 11, 1 (2021).

[70] H. H. Rashidi, S. Sen, T. L. Palmieri, T. Blackmon, J. Wajda, and N. K. Tran, Early recognition of burn-and trauma-related acute kidney injury: a pilot comparison of machine learning techniques, Scientific reports 10, 1 (2020).

[71] A. Mincholé and B. Rodriguez, Artificial intelligence for the electrocardiogram, Nature medicine 25, 22 (2019).

[72] Y. Tolkach, T. Dohmgörgen, M. Toma, and G. Kristiansen, High-accuracy prostate cancer pathology using deep learning, Nature Machine Intelligence 2, 411 (2020).

[73] J. Dukart, S. Weis, S. Genon, and S. B. Eickhoff, Towards increasing the clinical applicability of machine learning biomarkers in psychiatry, Nature Human Behaviour 5, 431 (2021).

[74] R. C. Li, S. M. Asch, and N. H. Shah, Developing a delivery science for artificial intelligence in healthcare, NPJ digital medicine 3, 1 (2020).

[75] Y. Park and M. Kellis, Deep learning for regulatory genomics, Nature biotechnology 33, 825 (2015).

[76] Y. Ito, M. Unagami, F. Yamabe, Y. Mitsui, K. Nakajima, K. Nagao, and H. Kobayashi, A method for utilizing automated machine learning for histopathological classification of testis based on johnsen scores, Scientific reports 11, 1 (2021).

[77] J. Kim, J. Lee, E. Park, and J. Han, A deep learning model for detecting mental illness from user content on social media, Scientific reports 10, 1 (2020).

[78] Y. Li, C. M. Nowak, U. Pham, K. Nguyen, and L. Bleris, Cell morphology-based machine learning models for human cell state classification, NPJ systems biology and applications 7, 1 (2021).

[79] X. Yu, W. Pang, Q. Xu, and M. Liang, Mammographic image classification with deep fusion learning, Scientific Reports 10, 1 (2020).

[80] M. Bracher-Smith, K. Crawford, and V. Escott-Price, Machine learning for genetic prediction of psychiatric disorders: a systematic review, Molecular Psychiatry 26, 70 (2021).

[81] D. Patel, V. Kher, B. Desai, X. Lei, S. Cen, N. Nanda, A. Gholamrezanezhad, V. Duddalwar, B. Varghese, and A. A. Oberai, Machine learning based predictors for covid-19 disease severity, Scientific Reports 11, 1 (2021).

[82] M. E. Newman, The structure and function of complex networks, SIAM review 45, 167 (2003).

[83] M. E. Newman, Assortative mixing in networks, Physical review letters 89, 208701 (2002).

[84] R. Albert and A.-L. Barabási, Statistical mechanics of complex networks, Reviews of modern physics 74, 47 (2002).

[85] L. C. Freeman, A set of measures of centrality based on betweenness, Sociometry, 35 (1977).

[86] L. C. Freeman, Centrality in social networks conceptual clarification, Social networks 1, 215 (1978).

[87] P. Bonacich, Power and centrality: A family of measures, American journal of sociology 92, 1170 (1987).

[88] R. Albert, H. Jeong, and A.-L. Barabási, Diameter of the world-wide web, nature 401, 130 (1999).

[89] J. M. Kleinberg, Hubs, authorities, and communities, ACM computing surveys (CSUR) 31, 5 (1999).

[90] D. Eppstein, M. S. Paterson, and F. F. Yao, On nearest-neighbor graphs, Discrete & Computational Geometry 17, 263 (1997).

[91] J. Doyle and J. Graver, Mean distance in a graph, Discrete Mathematics 17, 147 (1977).

[92] t. A. Snijders, The degree variance: an index of graph heterogeneity, Social networks 3, 163 (1981).

[93] M. Dehmer and A. Mowshowitz, A history of graph entropy measures, Information Sciences 181, 57 (2011).

[94] D. J. Watts and S. H. Strogatz, Collective dynamics of ‘small-world’networks, Nature 393, 440 (1998).

[95] M. E. Newman, D. J. Watts, and S. H. Strogatz, Random graph models of social networks, Proceedings of the National Academy of Sciences 99, 2566 (2002).

[96] S. B. Seidman, Network structure and minimum degree, Social networks 5, 269 (1983).

[97] M. Newman, Networks: an introduction (Oxford university press, 2010).

[98] P. Hage and F. Harary, Eccentricity and centrality in networks, Social networks 17, 57 (1995).

[99] B. S. Anderson, C. Butts, and K. Carley, The interaction of size and density with graph-level indices, Social networks 21, 239 (1999).

[100] V. Latora and M. Marchiori, Economic small-world behavior in weighted networks, The European Physical Journal B-Condensed Matter and Complex Systems 32, 249 (2003).

[101] M. E. Newman, Communities, modules and large-scale structure in networks, Nature physics 8, 25 (2012).

[102] J. Kim and J.-G. Lee, Community detection in multilayer graphs: A survey, ACM SIGMOD Record 44, 37 (2015).

[103] X. Zhao, J. Liang, and J. Wang, A community detection algorithm based on graph compression for large-scale social networks, Information Sciences 551, 358 (2021).

[104] A. Clauset, M. E. Newman, and C. Moore, Finding community structure in very large networks, Physical review E 70, 066111 (2004).

[105] M. Rosvall, D. Axelsson, and C. T. Bergstrom, The map equation, The European Physical Journal Special Topics 178, 13 (2009).

[106] M. E. Newman, Finding community structure in networks using the eigenvectors of matrices, Physical review E 74, 036104 (2006).

[107] U. N. Raghavan, R. Albert, and S. Kumara, Near linear time algorithm to detect community structures in large-scale networks, Physical review E 76, 036106 (2007).

[108] M. Girvan and M. E. Newman, Community structure in social and biological networks, Proceedings of the national academy of sciences 99, 7821 (2002).

[109] J. Reichardt and S. Bornholdt, Statistical mechanics of community detection, Physical review E 74, 016110 (2006).

[110] V. D. Blondel, J.-L. Guillaume, R. Lambiotte, and E. Lefebvre, Fast unfolding of communities in large networks, Journal of statistical mechanics: theory and experiment 2008, P10008 (2008).

[111] Y. Tian, H. Zhang, W. Xu, H. Zhang, L. Yang, S. Zheng, and Y. Shi, Spectral entropy can predict changes of working memory performance reduced by short-time training in the delayed-match-to-sample task, Frontiers in human neuroscience 11, 437 (2017).

[112] A. L. Vanluchene, H. Vereecke, O. Thas, E. P. Mortier, S. L. Shafer, and M. M. Struys, Spectral entropy as an electroencephalographic measure of anesthetic drug effect: a comparison with bispectral index and processed midlatency auditory evoked response, The Journal of the American Society of Anesthesiologists 101, 34 (2004).

[113] t. Elbert, W. Lutzenberger, B. Rockstroh, P. Berg, and R. Cohen, Physical aspects of the eeg in schizophrenics, Biological psychiatry 32, 595 (1992).

[114] B. Hjorth, Time domain descriptors and their relation to a particular model for generation of eeg activity, CEAN-Computerized EEG analysis, 3 (1975).

[115] B. Hjorth, Physical aspects of eeg data as a basis for topographic mapping, Topographic mapping of brain electrical activity, 175 (1986).

[116] Y. Bai, Z. Liang, and X. Li, A permutation lempel-ziv complexity measure for eeg analysis, Biomedical Signal Processing and Control 19, 102 (2015).

[117] A. Lempel and J. Ziv, On the complexity of finite sequences, IEEE Transactions on information theory 22, 75 (1976).

[118] S. Raschka and V. Mirjalili, Python machine learning: Machine learning and deep learning with Python, scikitlearn, and TensorFlow 2 (Packt Publishing Ltd, 2019).

[119] M. Mijalkov, E. Kakaei, J. B. Pereira, E. Westman, G. Volpe, and A. D. N. Initiative, Braph: a graph theory software for the analysis of brain connectivity, PloS one 12, e0178798 (2017).

[120] C. M. Michel and D. Brunet, Eeg source imaging: a practical review of the analysis steps, Frontiers in neurology 10, 325 (2019).

[121] E. E. Asher, M. Plotnik, M. Günther, S. Moshel, O. Levy, S. Havlin, J. W. Kantelhardt, and R. P. Bartsch, Connectivity of eeg synchronization networks increases for parkinson’s disease patients with freezing of gait, Communications biology 4, 1 (2021).

[122] C. L. Scrivener and A. T. Reader, Variability of eeg electrode positions and their underlying brain regions: visualizing gel artifacts from a simultaneous eeg-fmri dataset, Brain and behavior 12, e2476 (2022).

[123] R. Leech, R. Braga, and D. J. Sharp, Echoes of the brain within the posterior cingulate cortex, Journal of Neuroscience 32, 215 (2012).

[124] R. Leech and D. J. Sharp, The role of the posterior cingulate cortex in cognition and disease, Brain 137, 12 (2014).

[125] C. Windischberger, C. Lamm, H. Bauer, and E. Moser, Human motor cortex activity during mental rotation, NeuroImage 20, 225 (2003).

[126] J. Yang and H. Shu, The role of the premotor cortex and the primary motor cortex in action verb comprehension: Evidence from granger causality analysis, Brain research bulletin 88, 460 (2012).

[127] H. Duffau, L. Capelle, D. Denvil, P. Gatignol, N. Sichez, M. Lopes, J.-P. Sichez, and R. Van Effenterre, The role of dominant premotor cortex in language: a study using intraoperative functional mapping in awake patients, Neuroimage 20, 1903 (2003).

[128] N. B. Mota, N. A. Vasconcelos, N. Lemos, A. C. Pieretti, O. Kinouchi, G. A. Cecchi, M. Copelli, and S. Ribeiro, Speech graphs provide a quantitative measure of thought disorder in psychosis, PloS one 7, e34928 (2012).

[129] Q. Welniarz, C. Gallea, J.-C. Lamy, A. Méneret, T. Popa, R. Valabregue, B. Béranger, V. Brochard, C. Flamand-Roze, O. Trouillard, et al., The supplementary motor area modulates interhemispheric interactions during movement preparation, Human brain mapping 40, 2125 (2019).

[130] S. Walther, L. Schäppi, A. Federspiel, S. Bohlhalter, R. Wiest, W. Strik, and K. Stegmayer, Resting-state hyperperfusion of the supplementary motor area in catatonia, Schizophrenia bulletin 43, 972 (2017).

[131] M. L. Seghier, The angular gyrus: multiple functions and multiple subdivisions, The Neuroscientist 19, 43 (2013).

[132] O. Felician, P. Romaiguère, J.-L. Anton, B. Nazarian, M. Roth, M. Poncet, and J.-P. Roll, The role of human left superior parietal lobule in body part localization, Annals of Neurology: Official Journal of the American Neurological Association and the Child Neurology Society 55, 749 (2004).

[133] M. Koenigs, A. K. Barbey, B. R. Postle, and J. Grafman, Superior parietal cortex is critical for the manipulation of information in working memory, Journal of Neuroscience 29, 14980 (2009).

[134] S. Bhattacharjee, R. Kashyap, T. Abualait, S.-H. Annabel Chen, W.-K. Yoo, and S. Bashir, The role of primary motor cortex: more than movement execution, Journal of Motor Behavior 53, 258 (2021).

[135] A. Patel, G. M. N. R. Biso, and J. B. Fowler, Neuroanatomy, temporal lobe, in StatPearls [Internet] (StatPearls Publishing, 2021).

[136] A. Sritharan, P. Line, A. Sergejew, R. Silberstein, G. Egan, and D. Copolov, Eeg coherence measures during auditory hallucinations in schizophrenia, Psychiatry research 136, 189 (2005).

[137] K. R. Henshall, A. A. Sergejew, G. Rance, C. M. McKay, and D. L. Copolov, Interhemispheric eeg coherence is reduced in auditory cortical regions in schizophrenia patients with auditory hallucinations, International journal of psychophysiology 89, 63 (2013).

[138] S. Ryu, H. Lee, D.-K. Lee, H. J. Nam, Y.-C. Chung, and S.-W. Kim, Network structures of social functioning domains in schizophrenia and bipolar disorder: A preliminary study, Clinical Psychopharmacology and Neuroscience 18, 571 (2020).

[139] B. J. Duff, K. A. Macritchie, T. W. Moorhead, S. M. Lawrie, and D. H. Blackwood, Human brain imaging studies of disc1 in schizophrenia, bipolar disorder and depression: a systematic review, Schizophrenia research 147, 1 (2013).

[140] J. Gomez-Pilar, R. de Luis-García, A. Lubeiro, N. de Uribe, J. Poza, P. Núñez, M. Ayuso, R. Hornero, and V. Molina, Deficits of entropy modulation in schizophrenia are predicted by functional connectivity strength in the theta band and structural clustering, NeuroImage: Clinical 18, 382 (2018).

[141] t. Takahashi, R. Y. Cho, T. Mizuno, M. Kikuchi, T. Murata, K. Takahashi, and Y. Wada, Antipsychotics reverse abnormal eeg complexity in drug-naive schizophrenia: a multiscale entropy analysis, Neuroimage 51, 173 (2010).

[142] V. Molina, A. Lubeiro, R. de Luis Garcia, J. Gomez-Pilar, O. Martín-Santiago, M. Iglesias-Tejedor, P. Holgado-Madera, R. Segarra-Echeverría, M. Recio-Barbero, P. Núñez, et al., Deficits of entropy modulation of the eeg: A biomarker for altered function in schizophrenia and bipolar disorder?, Journal of Psychiatry and Neuroscience 45, 322 (2020).

[143] B. Hjorth, Eeg analysis based on time domain properties, Electroencephalography and clinical neurophysiology 29, 306 (1970).

[144] G. V. Portnova and M. S. Atanov, Nonlinear eeg parameters of emotional perception in patients with moderate traumatic brain injury, coma, stroke and schizophrenia, AIMS neuroscience 5, 221 (2018).

[145] D. Rangaprakash, M. N. Dretsch, J. S. Katz, T. S. Denney Jr, and G. Deshpande, Dynamics of segregation and integration in directional brain networks: illustration in soldiers with ptsd and neurotrauma, Frontiers in neuroscience, 803 (2019).

[146] W. Luo, A. S. Greene, and R. T. Constable, Within node connectivity changes, not simply edge changes, influence graph theory measures in functional connectivity studies of the brain, NeuroImage 240, 118332 (2021).

[147] D. S. Bassett, E. Bullmore, B. A. Verchinski, V. S. Mattay, D. R. Weinberger, and A. Meyer-Lindenberg, Hierarchical organization of human cortical networks in health and schizophrenia, Journal of Neuroscience 28, 9239 (2008).

[148] J. Golbeck, Analyzing the social web (Newnes, 2013).

[149] J. B. Thomas, M. R. Brier, M. Ortega, T. L. Benzinger, and B. M. Ances, Weighted brain networks in disease: centrality and entropy in human immunodeficiency virus and aging, Neurobiology of aging 36, 401 (2015).

[150] A. Griffa, P. S. Baumann, C. Ferrari, K. Q. Do, P. Conus, J.-P. Thiran, and P. Hagmann, Characterizing the connectome in schizophrenia with diffusion spectrum imaging, Human brain mapping 36, 354 (2015).

[151] C. A. Baltazar, B. S. Machado, D. D. De Faria, A. J. M. Paulo, S. M. C. A. Silva, H. B. Ferraz, and P. de Carvalho Aguiar, Brain connectivity in patients with dystonia during motor tasks, Journal of Neural Engineering 17, 056039 (2020).

[152] I. Goodfellow, Y. Bengio, and A. Courville, Deep learning (MIT press, 016).

[153] N. Srivastava, G. Hinton, A. Krizhevsky, I. Sutskever, and R. Salakhutdinov, Dropout: a simple way to prevent neural networks from overfitting, The journal of machine learning research 15, 1929 (2014).

